# The 2020 SARS-CoV-2 epidemic in England: key epidemiological drivers and impact of interventions

**DOI:** 10.1101/2021.01.11.21249564

**Authors:** Edward S. Knock, Lilith K. Whittles, John A. Lees, Pablo N. Perez-Guzman, Robert Verity, Richard G. FitzJohn, Katy AM Gaythorpe, Natsuko Imai, Wes Hinsley, Lucy C. Okell, Alicia Rosello, Nikolas Kantas, Caroline E. Walters, Sangeeta Bhatia, Oliver J Watson, Charlie Whittaker, Lorenzo Cattarino, Adhiratha Boonyasiri, Bimandra A. Djaafara, Keith Fraser, Han Fu, Haowei Wang, Xiaoyue Xi, Christl A. Donnelly, Elita Jauneikaite, Daniel J. Laydon, Peter J White, Azra C. Ghani, Neil M. Ferguson, Anne Cori, Marc Baguelin

**Affiliations:** MRC Centre for Global Infectious Disease Analysis, Abdul Latif Jameel Institute for Disease and Emergency Analytics (J-IDEA), School of Public Health, Imperial College London; UK; National Institute for Health Research Health Protection Research Unit in Modelling and Health Economics, UK; Department of Infectious Disease, School of Public Health, Imperial College London; UK; Department of Infectious Disease Epidemiology, Faculty of Epidemiology and Population Health, London School of Hygiene and Tropical Medicine, London, UK; Faculty of Natural Sciences, Department of Mathematics, Imperial College London, UK; Department of Statistics, University of Oxford, Oxford, UK

## Abstract

We fitted a model of SARS-CoV-2 transmission in care homes and the community to regional surveillance data for England. Among control measures implemented, only national lockdown brought the reproduction number below 1 consistently; introduced one week earlier it could have reduced first wave deaths from 36,700 to 15,700 (95%CrI: 8,900–26,800). Improved clinical care reduced the infection fatality ratio from 1.25% (95%CrI: 1.18%–1.33%) to 0.77% (95%CrI: 0.71%–0.84%). The infection fatality ratio was higher in the elderly residing in care homes (35.9%, 95%CrI: 29.1%–43.4%) than those residing in the community (10.4%, 95%CrI: 9.1%–11.5%). England is still far from herd immunity, with regional cumulative infection incidence to 1st December 2020 between 4.8% (95%CrI: 4.4%–5.1%) and 15.4% (95%CrI: 14.9%–15.9%) of the population.

**One-sentence summary:** We fit a mathematical model of SARS-CoV-2 transmission to surveillance data from England, to estimate transmissibility, severity, and the impact of interventions

## 1 Introduction

England is among the countries worst-affected by the global pandemic of COVID-19, caused by the novel *Betacoronavirus* SARS-CoV-2. As of 2^nd^ December 2020, over 51,000 deaths have been reported nationally, or 91 deaths per 100,000 people (*1*). The impact of the epidemic has varied across the country, with regional epidemics differing in their severity and timing. A key feature in all regions is the burden suffered by older adults living in care homes, where mortality has been high.

We use a mathematical model of SARS-CoV-2 transmission to reproduce the first two waves of the epidemic across England’s seven NHS regions and assess the impact of interventions implemented by the UK government. We analyse the epidemic from importation of SARS-CoV-2 into each region to the 2^nd^ December 2020: encompassing the first national lockdown from March –May, the interventions implemented as COVID-19 deaths increased again in the autumn, eventually leading to the second national lockdown in November.

We built an age-structured stochastic transmission model of SARS-CoV-2, representing care homes, hospital clinical pathways and the wider community (Materials and Methods). We developed a Bayesian evidence-synthesis approach to estimate model parameters and to reconstruct regional epidemics using data from daily recorded deaths, PCR testing, hospital admissions, hospital bed occupancy, individual patient outcomes, contact surveys, and serological surveys. We evaluated temporal changes in transmission as new control measures were implemented and then relaxed, and population immunity accrued. Inclusion of serological data allowed us to robustly estimate region- and age-specific disease severity, to compare severity in care home residents to elderly individuals in the community, and estimate the total epidemic size, by calculating the proportion of individuals infected over time in each region. Finally, we examined counterfactual epidemic scenarios, varying the date and duration of the first national lockdown and the effectiveness of restricting care home visits, to quantify the resulting impact on mortality.

Our analysis, which synthesises multiple data sources and parametrically accounts for their biases, provides a comprehensive overview of transmission, hospitalisation, and mortality patterns of SARS-CoV-2 in the first and second waves (up to 2^nd^ December) in all regions of England. Our results provide crucial insights for controlling the epidemic in the future, emphasising the importance of acting fast to save lives.

## 2 Results

### 2.1 Epidemic trajectory

We used our evidence-synthesis approach, to infer the COVID-19 epidemic start date in each NHS England region (Figure 1A), then reconstructed epidemic trajectories for hospitalisations (Figure S7) and deaths in care homes and hospitals (Figure 1B-H). We estimated the basic reproduction number, *R*_*0*_, defined as the expected number of onward infections from an infectious individual in a fully susceptible population to be 2.9 (95% CrI: 2.8-3.1) nationally. Figure 1I shows how the effective reproduction number 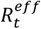 (the expected number of onward infections from an individual infected at time t) changed in each region over time, in relation to government control measures and accrual of population immunity.

**Figure 1:**
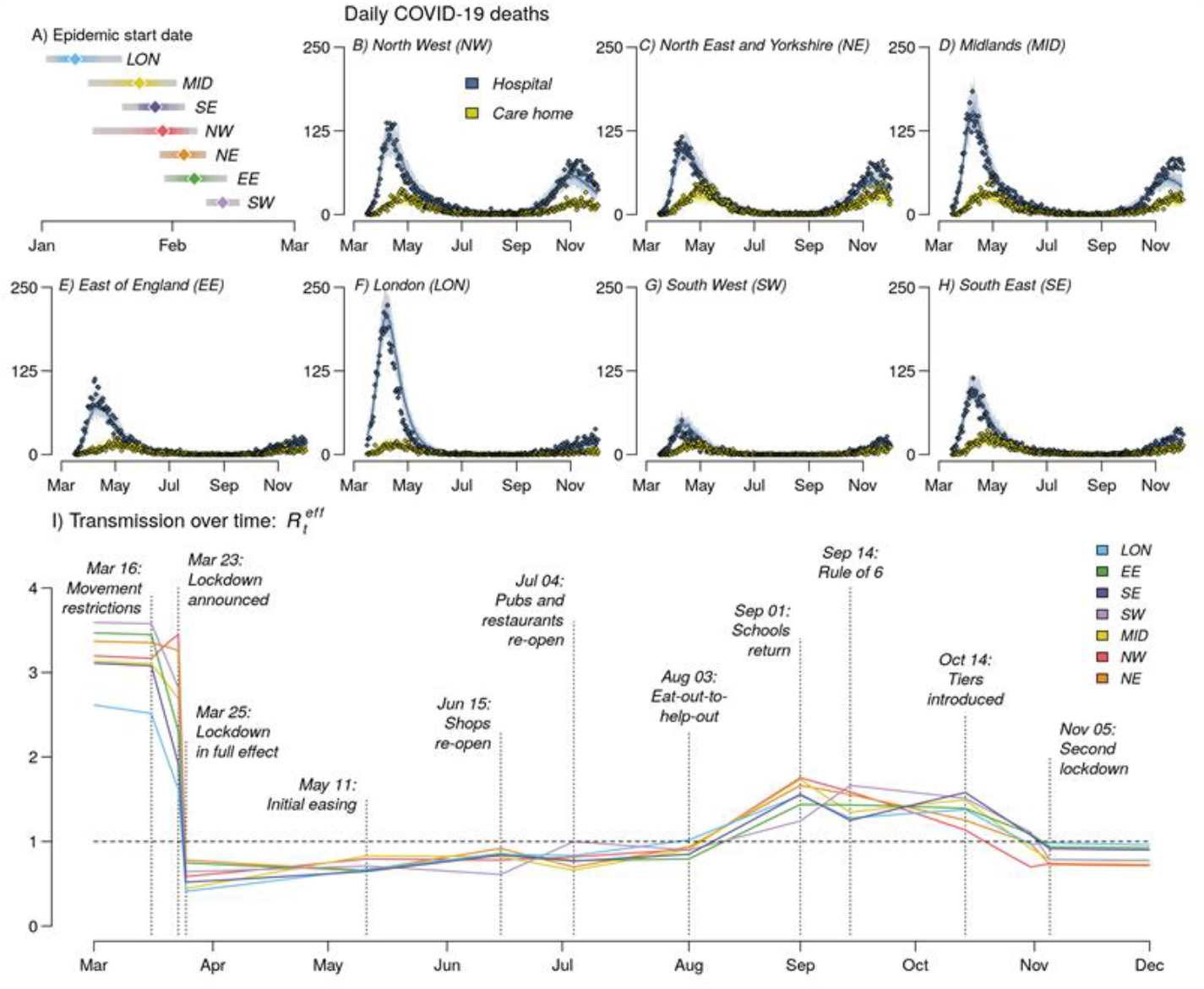
Trajectory of the England COVID-19 epidemic. **A**, The inferred epidemic start date in each NHS England region. **B-H**, The model fit to reported daily deaths from COVID-19 in care homes and hospitals for each NHS England region. The points show the daily data, solid lines the median posterior and the shaded area shows the 95% CrI. **I**, The mean effective reproduction number within the general community (i.e. excluding care homes) in each region from March to December. Vertical lines and labels represent dates of key policy changes, defining the breaking points of the underlying piecewise linear transmission rate. Dashed horizontal line depicts reproduction number of 1.

The first COVID-19 death in England occurred on 5^th^ March 2020 (*2*). Seven days later, in response to the growing epidemic, the government began to introduce control measures, initially requiring individuals with a dry persistent cough and/or fever to self-isolate (*3*). On 23^rd^ March this escalated to a full national lockdown (*4, 5*). Irrespective of initial differences, the level of transmission during lockdown was similar across all regions (Figure 1I), consistent with mobility data showing movement during lockdown reduced to a consistent level nationally (*6*).

The epidemic in London began 15 days before (95% CrI: 28 days before, 3 days after) the rest of the country (Figure 1A), meaning the lockdown occurred at a later stage of its epidemic. London experienced a mortality of 88.5 (95% CrI: 79.9–95.3) per 100,000 during the first wave, compared to the national average of 70.7 (95% CrI: 64.6–77.1), despite having a younger population and a smaller care home population than other regions (296 vs 603 per 100,000 nationally).

A key feature of the first epidemic wave in England, in common with other European countries, was the high death toll within care homes, which accounted for 22.6% laboratory-confirmed COVID-19 deaths in England as of 1^st^ August 2020. Although community transmission rates fell during lockdown, transmission within care homes continued to rise, with infection risk peaking in care home residents, between 26^th^ March in London and 12^th^ April in North East and Yorkshire (Figure 2A). Deaths in care homes peaked on average 13 days later than hospital deaths (Figure 1B-H).

**Figure 2:**
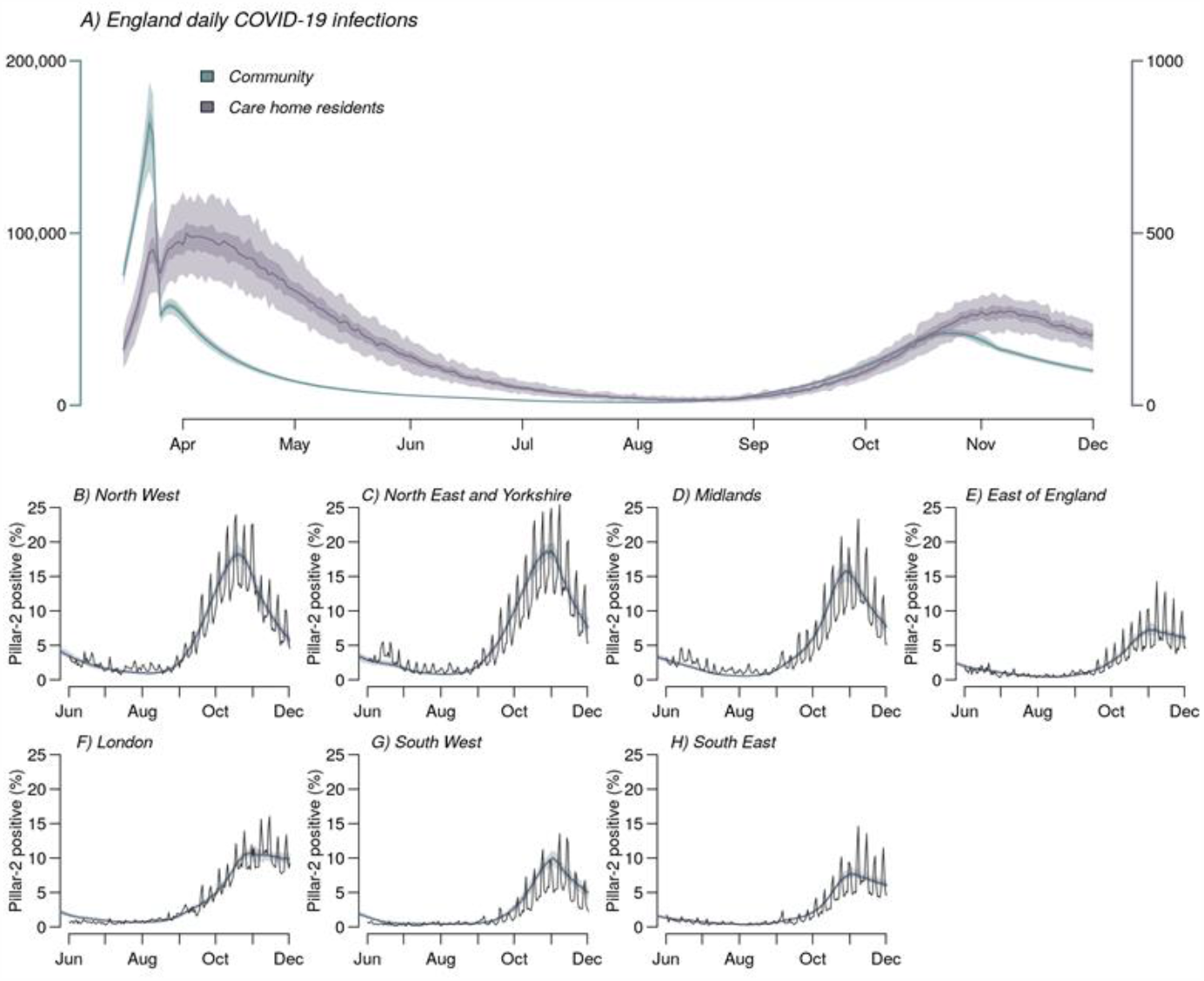
**A**, Inferred daily SARS-CoV-2 infections in England care home residents (right axis) and the wider community (left axis). **B-H**, Comparison of modelled (shaded bands) and observed (solid line) proportion of PCR tests that are positive, under pillar-2 testing (community swab testing for symptomatic individuals) in >25 year olds. Shaded bands depict 95% CrI, 50% CrI and median model outputs.

The first lockdown in England continued until 11^th^ May, when people unable to work remotely were permitted to resume their jobs. Over the summer restrictions were successively eased, with non-essential shops, pubs and restaurants opening, followed by the government’s ‘Eat Out to Help Out’ restaurant subsidy scheme in August (*7*). This led to a steady increase in transmission, with 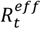 rising above 1 in all regions by mid-August (Figure 1I).

Increasing PCR test positivity marked the beginning of a second epidemic wave (Figure 2B-H, S6). The accompanying introduction of non-pharmaceutical interventions (NPIs) began with the “Rule of Six” (limiting social gatherings to 6 persons maximum) on 14^th^ September (*8*), followed by the localised tiered restrictions on 14^th^ October (*9*). These measures limited transmission in most regions but were not sufficient to reduce 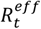 below 1 (Figure 1I). Consequently, on 31^st^ October, the government announced a second national lockdown, which lasted from 5^th^ November to 1^st^ December (*10*).

Restrictions during the second lockdown were less stringent than the first, with schools and some workplaces remaining open. This was reflected in 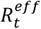 estimates of 0.83 (95% CrI: 0.81–0.85) at the start of the second lockdown, compared to 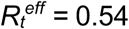 (95% CrI: 0.50–0.59) at the start of the first. We estimate that without the population immunity accrued during the first wave, contact rates during the second lockdown would have resulted in a reproduction number of *R*_*t*_ *=* 0.95 (95% CrI: 0.93–0.98). Hence, population immunity helped to reduce transmission further below the critical threshold of 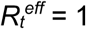.

### 2.2 Severity and hospitalisation

COVID-19 manifests a broad spectrum of severity, from asymptomatic infection to life-threatening illness requiring intensive care. We estimated age-patterns of clinical progression in people admitted to hospital using individual-level data from 17,702 patients admitted between 18^th^ March and 31^st^ May 2020 (inclusive) in the COVID-19 Hospitalisation in England Surveillance System (CHESS, (*11*)) (Materials and Methods). We derived estimates of the time spent in each stage of the hospital pathway (including general wards, ICU and post-ICU stepdown care), as well as age-stratified probabilities of progression through that pathway (Figure 3 and Figure S8). We accounted for differing length of stays given different outcomes; there were marked differences in average length of ICU stay for those who died in ICU, those who later died in stepdown care and those who were discharged following stepdown care (Figure 3F). Among patients over 65, we found the probability of admission to ICU decreased with increasing age. Severity of COVID-19 increases with age, but for older patients and those with most severe illness, the benefit of ICU admission, ventilation and the corresponding prognosis may not be better than with oxygen therapy in a general ward (*12*). Thus, older and more severely infected patients may be directed to care on a general ward rather than admitted to ICU.

**Figure 3:**
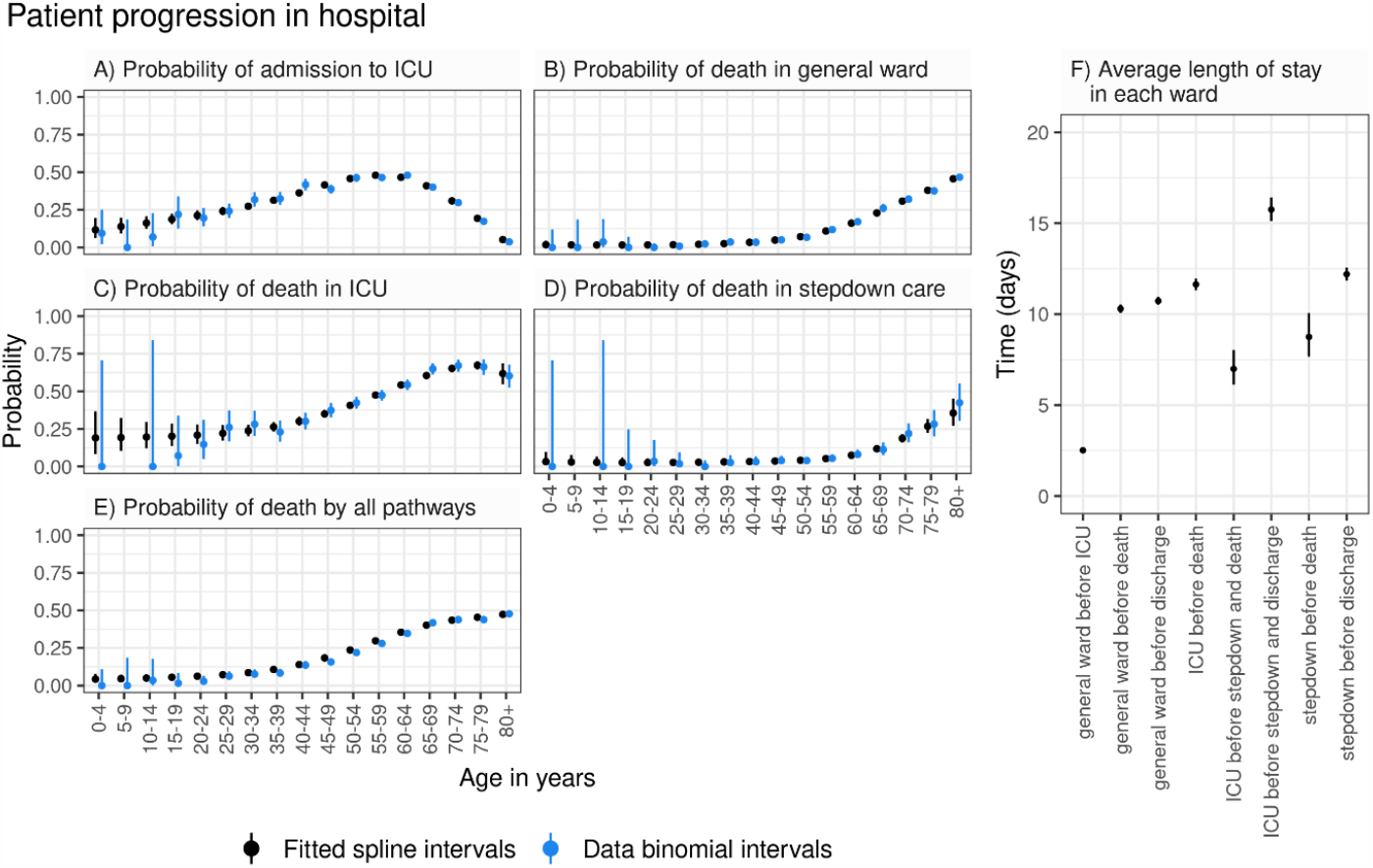
Age-dependent probabilities of progression through hospital pathways. **A**, Probability of admission to ICU. **B**, Probability of death in a general ward. **C**, Probability of death in ICU. **D**, Probability of death in stepdown care. **E**, Probability of death through all hospital pathways. Black circles and vertical segments show posterior mean and 95% credible intervals of splines fitted to data, blue circles and vertical segments show raw mean values and 95% confidence intervals (exact binomial) for each 5-year age group. **F**, Average length of stay in each ward (posterior mean and 95% credible intervals).

We used estimates of clinical progression to parametrise the transmission model, enabling us to infer temporal and regional differences in disease severity, informed by local demography, observed daily hospital admissions, bed occupancy and deaths. We measured severity of disease by the infection fatality ratio (IFR) and the infection hospitalisation ratio (IHR).

The severity of disease increased with age in all regions with the steepest increase above 65 years (Figure 4A-C), in line with observations worldwide (5). Regional estimates of age-aggregated disease severity depend on the population age distribution, which is similar in most regions of the country, except London, where the median age is 34.6 years (vs 39.5 years nationally). At the start of the first wave, London experienced an IFR (respectively IHR) of 0.91% (95% CrI: 0.82%–1.00%) (resp. 3.02%; 95% CrI: 2.82%–3.19%) compared to the national average of 1.25% (95% CrI: 1.18%–1.33%) (resp. 3.52%; 95% CrI: 3.29%–3.72%) (Figure 4D-E).

**Figure 4:**
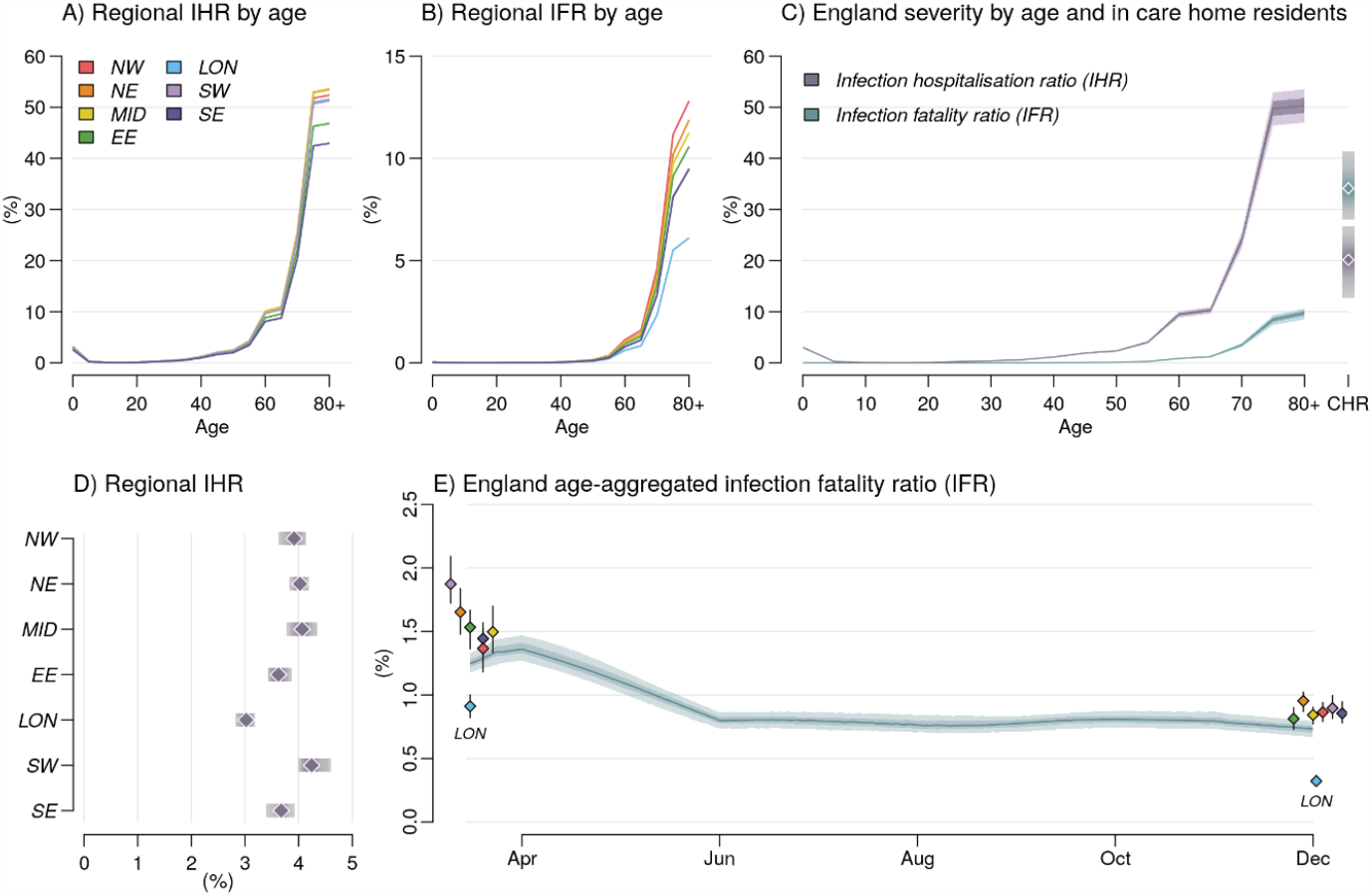
Relative severity of disease by age group and region. **A**, and **B**, Variation in the Infection fatality ratio (IFR) and Infection Hospitalisation Ratio (IHR) by age group in each region. Ages 80+ were modelled as a single risk group, care home residents are not included. **C**, The England IFR and IHR by age group and in care home residents (estimates denoted CHR at the right-hand side of the panel). National severity estimates are produced by aggregating regional estimates based on infection incidence. **D**, The regional IHR, aggregated over age and risk group by infection incidence. Plots a-d use parameter estimates, and incidence weightings calculated as of 1^st^ December 2020. **E**, The England IFR over time, coloured dots show regional estimates of IFR at the start of the epidemic and on 1^st^ December 2020 (clusters each correspond to one time-point, LON: London). In plots **C-E** Shaded bands depict 95% CrI and interquartile ranges, points depict medians.

Regional variation in the population age distribution did not fully account for differences in severity, with London still experiencing lower mortality when stratified by age (Figure 4A-B). The oldest age group (80+) in London had an IFR of 6.1% (95% CrI: 5.2%–6.8%) compared to 12.7% (95% CrI: 10.8%–14.3%) in the North West.

We estimated temporal trends in the IFR for England, by weighting regional estimates by incidence and population demographics. At the start of the first wave, the national IFR was 1.25% (95% CrI: 1.18%–1.33%) (Figure 4E), consistent with earlier reports from serology data alone (*13*). The national IFR initially appeared to increase, as transmission widened from London to regions with older populations and greater disease severity. Over the first wave, the proportion of hospital admissions resulting in death decreased, due to improvements in clinical management and alleviation of capacity constraints (*14*), leading to a national IFR of 0.77% (95% CrI: 0.71%–0.84%) by the end of the first wave. The magnitude of the relative reduction in IFR over time varied between regions, from 36.5% (95% CrI: 26.5%–47.5%) in the North West to 64.6% (95% CrI: 58.6%–68.8%) in London.

The IFR was greater among care home residents (35.9%, 95% CrI: 29.1%–43.4%) than in the 80+ in the community (10.4%, 95% CrI: 9.1%–11.5%, Figure 4C). Many care home residents did not transfer into hospital, and instead died in the facilities where they lived, so conversely the IHR was lower in care home residents (19.1%, 95% CrI: 11.5%–26.8%) than in those aged 80+ in the community (51.1%, 95% CrI: 47.6%–54.3%). We present national estimates of severity at the end of the second wave, stratified by age and care home residency in Table S9.

### 2.3 Epidemic size

Data from repeated serological surveys of blood donors aged 15-65 informed our estimation of the total regional epidemic size (Figure 5A-G), accounting for imperfect sensitivity and specificity of serological tests (Materials and Methods) (*15*). The cumulative proportion of the population ever infected with SARS-CoV-2 ranged from 4.8% (95% CrI: 4.4%–5.1%) in the South West to 15.4% (95% CrI: 14.9%–15.9%) in London (Figure 5H). Predicted seropositivity was initially greater than cumulative incidence, due to imperfect test specificity. The increase in seropositivity lagged cumulative infections by two weeks, reflecting the time from infection to seroconversion.

**Figure 5:**
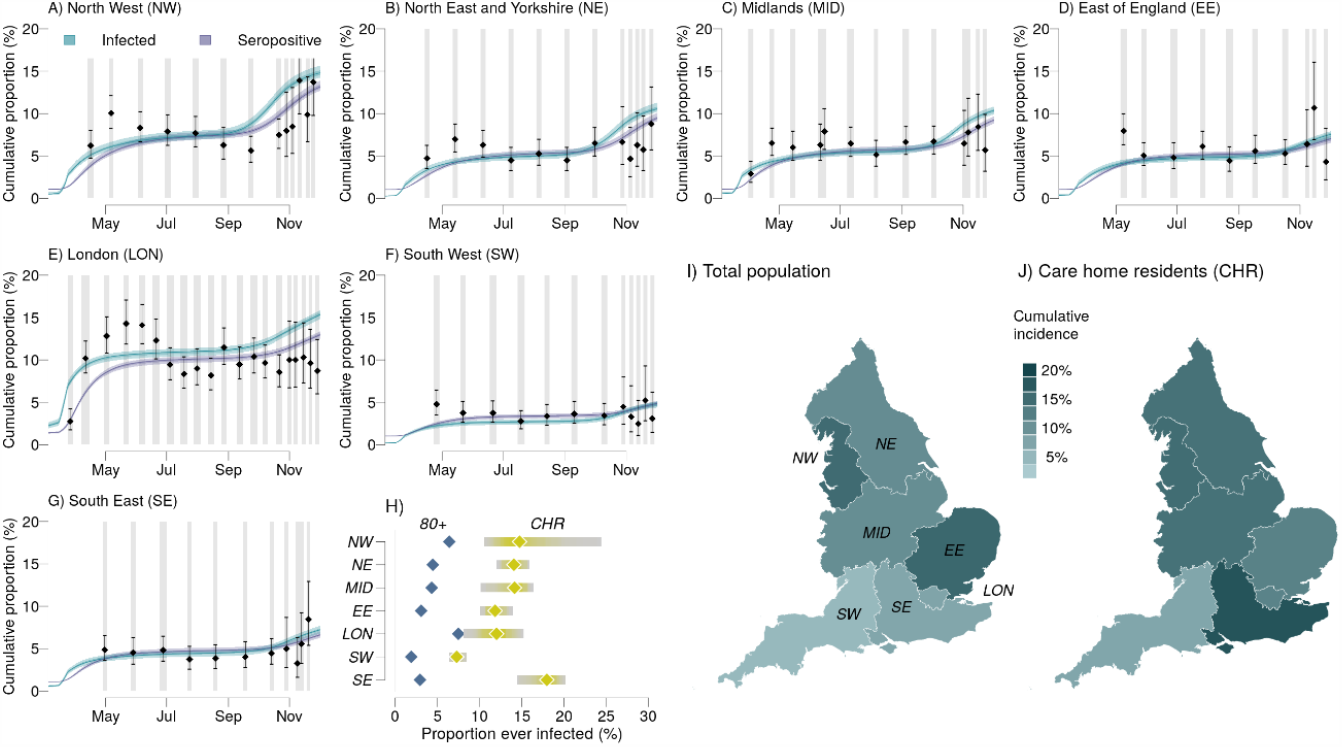
Cumulative incidence and seropositivity by region. **A-G**, Comparison of the estimated proportion of the population testing seropositive with observations from serological surveys. Vertical grey shaded bands show serological survey timings, black points the observed seroprevalence (bars: 95% exact confidence intervals), blue and purple lines show estimated proportion of the population infected and seropositive respectively (shaded bands the 95% CrI, 50% CrI and median). **H**, Comparison by region of the estimated cumulative attack rate in care home residents vs in the 80+ age group in the community (median, 95% CrI). The final epidemic size in each England NHS region **I**) in total and **J**) in care home residents.

Seropositivity notably declined following the first wave in some regions (Figure 5A-G). This may reflect antibody waning (*16*), or temporal trends in the composition of the surveyed population. Lockdown restrictions made attending blood donation centres difficult for all except key workers, who were more likely to have been infected (*17*), and may therefore be overrepresented in the sample of blood donors during the two lockdowns.

The proportion of care home residents ever infected with SARS-CoV-2 was 13.7% (95% CrI: 10.7%–16.7%), much higher than the 4.2% (95% CrI: 4.0%–4.4%) estimated in >80-year olds living in the community. This difference was consistently observed across all regions (Figure 5H). Regional differences in care home attack rates mirrored the patterns seen in the general community, with regions with larger community epidemics also experiencing larger care home epidemics (Figure 5I,J).

### 2.4 Impact of non-pharmaceutical interventions (NPIs)

We explored counterfactual intervention scenarios and examined the potential impact on mortality of initiating the first national lockdown one week earlier or later; ending that lockdown two weeks earlier or later; and 50% more or less restricted care home visits throughout the epidemic (Figure 6).

**Figure 6:**
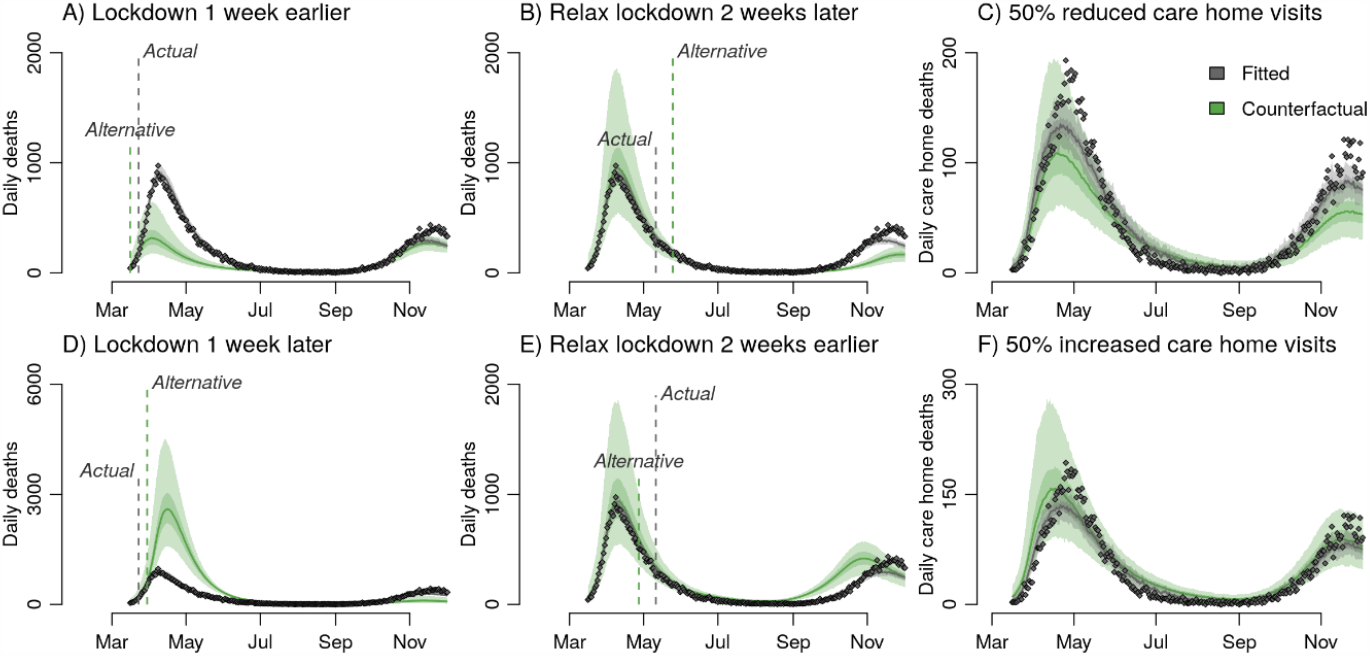
Counterfactual analysis of the impact on mortality aggregated across NHS England regions of **A, D**, initiating lockdown one week earlier / later, **B, E** Relaxing lockdown two weeks earlier / later, and **C, F** 50% more / less restricted care home visits. Panels **A, B, D** and **E** all present counterfactual outcomes for daily deaths in England but have different y-axis scales to better highlight differences between the observed data and each alternative lockdown scenario.

The timing of the initial national lockdown was crucial in determining the eventual epidemic size in England. Locking down a week earlier could have reduced the first wave death toll (up to 1^st^ July 2020) from 36,700 to 15,700 (95% CrI: 8,900–26,800) while delaying lockdown by a week would have increased the deaths to 102,600 (95% CrI: 66,400–154,800) (Figure 6A, D). The impact varied by region, with regions with less established epidemics at the time of the first lockdown more sensitive to the timing of the intervention (Figure S10). Locking down a week later may have increased deaths, with large variability by region, from 105% in London to 274% in the Midlands but with very large uncertainty (Figure S9). Initiating a lockdown to interrupt the exponential growth phase of an epidemic has a much greater impact on reducing total mortality than extending an existing lockdown. Due to this asymmetry, relaxing the lockdown measures two weeks earlier (respectively later), could have increased deaths by 9,300 (95% CrI: 700–17,000) (respectively prevented 9,800 (95% CrI: 7,400–12,100) deaths) prior to 2^nd^ December (Figure 6B, D).

We also explored counterfactual scenarios varying the level of visit restriction in care homes and estimated that reducing contact between the general population and care home residents by 50% could have reduced care home deaths by 44% (95% CrI: 17%–64%) (Figure 6C).

## 3 Discussion

We present a comprehensive overview of SARS-CoV-2 transmission, hospitalisation, mortality and intervention impact in the first two epidemic waves across all regions of England between March and December 2020. We successfully reproduce the transmission dynamics of the two epidemic waves, in terms of cases, PCR prevalence, seroprevalence, hospitalised cases (general wards and ICU), and deaths in hospitals and in care homes. We estimate intense transmission in care homes even during the first national lockdown when 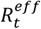 in the community was well below one in all regions (Figure 2) (*18*–*20*). Combined with our counterfactual analysis of restricting visits (Figure 6) this suggests that reducing infection levels in care home residents is challenging. This highlights the difficulty of protecting care home residents from COVID-19: due to the necessarily close contact between staff and residents within a care home, once a care home outbreak has begun it is very difficult to reduce transmission, which overrides any impact of reducing the number of introductions (*21, 22*).

We find that, consistent with existing literature (*23*), disease severity increases with age. Assessment of severity is complicated by the broad clinical spectrum of COVID-19 (*24*–*26*) hence, recent published estimates are still based on data from early in the pandemic (*27*). Here we provide updated severity estimates based on multiple contemporary data streams. We estimate considerable regional heterogeneity in severity, broadly consistent in the general population and in care homes for IFR and IHR. London experienced the lowest severity even after adjusting for its younger population. The estimated two-fold reduction over time in IFR (Figure 4) cannot be explained solely by the introduction of dexamethasone which reduces mortality amongst ICU patients (*28*), but rather a combination of factors including improvements in clinical management, greater experience in treating patients in ICU, and alleviation of capacity constraints (*14, 29*).

Our analysis shows large regional variation in burden, especially in the first wave. This is likely due to the pattern of seeding and the timing of lockdown relative to how advanced each region’s epidemic was (Figure 1A). Our counterfactual scenarios of initiating the first national lockdown one week earlier or later highlight the importance of early interventions to reduce overall mortality (Figure 6).

The extent and duration of infection-induced immunity to SARS-CoV-2 and its relationship to seropositivity remains unclear. Related seasonal coronaviruses induce immunity that wanes in one or two years (*30*), though antibody titres following SARS-CoV-1 infection appear to decay more slowly (*31*). Our estimated cumulative incidence over time (Figure 5), strongly supports the hypothesis that the epidemic decline after the first national lockdown was due to NPIs, with immunity playing a minimal role (*32*). Population-level immunity was insufficient to prevent a second wave of infection in any region (Figure 1), illustrated by the increase in reported cases and deaths which prompted the second national lockdown (*33*).

With the authorisation of the first SARS-CoV-2 vaccines in December 2020, we are now entering a new phase in the control of the COVID-19 pandemic. However, our estimates of current population immunity are low, with regional cumulative attack rates ranging from 4.8% to 15.4%, therefore any vaccination campaign will need to achieve high coverage and high levels of protection in vaccinated individuals to allow NPIs to be lifted without a resurgence of transmission. While vaccinating the most vulnerable age and risk groups will considerably reduce the burden of COVID-19, a large proportion of younger age groups may also need to be vaccinated to reach the immunity threshold for control. Our high estimates of transmission in care homes imply that vaccine uptake there will need to be especially high, particularly if vaccine efficacy is lower amongst older age groups.

We make a number of simplifying assumptions in our analysis. First, due to the compartmental nature of our model, we do not explicitly model individual care homes, rather the regional care home sector as a whole. However, as care home workers may work across multiple facilities leading to within and between care home transmission, we do not expect the simplification to substantially affect our conclusions. Similarly, we do not model individual households or transmission within and between them. When assessing the impact of NPIs on transmission we therefore capture population averages, rather than the contribution of household and non-household contacts. Second, hospital-acquired infections may have contributed to overall transmission, especially around the peak of the epidemic, and to persistence of infection in England over the summer months (*34, 35*). Our model does not explicitly represent nosocomial transmission; therefore such effects will be encompassed within our regional 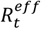 estimates. Third, each data stream was subject to competing biases, which we statistically accounted for as far as possible (supplement section 1.1.2). A key strength of our evidence-synthesis approach is that we do not rely on any single data source, combining multiple perspectives to provide a robust overall picture of the epidemic. Finally, we model the epidemics in each NHS region in England independently without accounting for spatial effects across regional boundaries, or spatial heterogeneity within regions. This spatial scale was determined by the data and reflects limited movement between regions due to travel restrictions but does allow for movement within regions.

Our analysis provides a comprehensive overview of transmission, hospitalisation, and mortality patterns of COVID-19 in the first and second waves of the epidemic in all regions of England, one of the countries worst-affected by the pandemic. Integration of multiple data streams into a single cohesive modelling framework, enables us to disentangle transmission and severity from features of the surveillance system and provide robust estimates of the epidemiological characteristics of the COVID-19 epidemic in England. As nationwide vaccination programmes are rolled out, our results will help to inform how NPIs are applied in the future.

## Data Availability

All code and de-identified regionally aggregated data (see supplementary materials for full details) required to reproduce this analysis are available at https://github.com/mrc-ide/sarscov2-transmission-england (https://zenodo.org/record/4384864)

https://zenodo.org/record/4384864

## Acknowledgements

We thank all the colleagues at Public Health England (PHE) and frontline health professionals who have driven and continue to drive the daily response to the epidemic, but also for providing the necessary data to inform this study. This work would not have been possible without their dedication and expertise. The use of pillar 2 PCR testing data was made possible thanks to PHE colleagues and we extend our thanks to Dr Nick Gent and Dr André Charlett for facilitation and their insights into these data. The use of serological data was made possible by colleagues at PHE Porton Down, Colindale, and the NHS Blood Transfusion Service. We are particularly grateful to Dr Gayatri Amirthalingam and Prof Nick Andrews for helpful discussions around these data. We also thank the entire Imperial College London Covid-19 Response Team for their support and feedback throughout. This work was supported by the NIHR HPRU in Modelling and Health Economics, a partnership between PHE, Imperial College London and LSHTM (grant code NIHR200908). We acknowledge funding from the MRC Centre for Global Infectious Disease Analysis (reference MR/R015600/1), jointly funded by the UK Medical Research Council (MRC) and the UK Foreign, Commonwealth & Development Office (FCDO), under the MRC/FCDO Concordat agreement and is also part of the EDCTP2 programme supported by the European Union.

## Disclaimer

The views expressed are those of the authors and not necessarily those of the United Kingdom (UK) Department of Health and Social Care, the National Health Service, the National Institute for Health Research (NIHR), Public Health England (PHE), UK MRC, UKRI or European Union.

## List of Supplementary Materials

Supplementary materials (Materials and Methods, Supplementary Results) Supplementary data files: data_rtm.csv, data_serology.csv, support_progression.csv, support_severity.csv

## Supplementary Materials

### 1 Materials and Methods

Understanding the transmission of SARS-CoV-2 is challenging. The available data are subject to competing biases, such as dependence on case definition for testing and reporting, as well as being influenced by capacity and logistical constraints. These factors are further complicated by the nature of SARS-CoV-2 transmission, whereby a substantial proportion of infected individuals develop very mild symptoms, or remain asymptomatic, but are nonetheless able to infect others (1). In this section, we describe the data used in our analyses, give details on the dynamic transmission model, and present the methods used for fitting the model to the various data sources, accounting for the inherent biases in those data.

#### 1.1 Data sources

Here we detail the datasets used to calibrate the model to the regional epidemics. We fitted our model to time series data spanning 16th March 2020 to 2nd December 2020 (inclusive), using the data available to us on 14th December 2020, by which point the effect of remaining reporting lags would be minimal.

##### 1.1.1 Hospital admissions and bed occupancy

We use healthcare data for each NHS region from the UK Government Dashboard (supplementary data files: *data_rtm.csv*, columns: *phe_admissions, phe_occupied, phe_patients*) (2).

For admissions data, we use the daily number of confirmed COVID-19 patients admitted to hospital, which includes people admitted to hospital who tested positive for COVID-19 in the 14 days prior to admission and inpatients who tested positive in hospital after admission, with the latter being reported as admitted on the day prior to their diagnosis.

For ICU bed occupancy, we use the daily number of (confirmed) COVID-19 patients in beds which are capable of delivering mechanical ventilation.

For the occupancy in general (i.e. non-ICU) hospital beds, we use the daily number of confirmed COVID-19 patients in hospital beds with ICU occupancy subtracted.

##### 1.1.2 Deaths

We use the number of deaths by date of death for people who had a positive COVID-19 test result and died within 28 days of their first positive test provided Public Health England.

These can be found on (2). We also use the number among these deaths occurring in hospital (as reported by NHS England) and consider the remainder to have occurred in care homes. While non-hospital deaths may include deaths in other settings, such as in private residences, comparison with ONS data suggests that care home deaths from COVID-19 may also have been under-reported. As such we consider non-hospital deaths to be an appropriate proxy for care home deaths, and do not expect the margin for under or over-ascertainment to affect our conclusions. These data were provided by PHE and the data we have been using is provided as a supplementary file (supplementary data file: *data_rtm.csv*, columns: *death2, death3*) to allow reproducibility of our analysis.

##### 1.1.3 Pillar 2 testing

We use pillar 2 testing data (see supplementary data files), which covers PCR testing for the general population (as compared with pillar 1 testing, which mainly occurred in hospitals). Since such testing was not available to the whole population for much of the spring wave of the pandemic, we only use this data from June 1^st^ onwards.

We use the daily number of positives and negative tests by specimen date. Each individual who tested positive was only counted once in the number of positives, on the specimen date of their first positive test. Multiple negatives were allowed per individual, but the negatives of all individuals who ever tested positive had been removed. We only consider PCR tests and thus exclude lateral flow tests, which have been introduced recently in trials of population mass testing. We also only use pillar 2 data for those aged 25 or over, to avoid bias resulting from increased testing of university students around the reopening of universities (supplementary data file: *data_rtm*.csv, columns: *pillar2_negatives_non_lft_over25, pillar2_positives_over25*).

##### 1.1.4 Serology surveys

Serological survey data come from antibody testing by Public Health England of samples from healthy adult blood donors, supplied by NHS Blood and Transplant (NHSBT) (supplementary data file: *data_serology.csv*).

##### 1.1.5 REACT-1 prevalence survey

We use the daily number of positives and negatives by specimen date from the first 7 rounds of the REACT-1 (Real-time Assessment of Community Transmission) infection prevalence survey (supplementary data file: *data_rtm.csv*, columns: *react_positive, react_samples*) (3). Note that results published in REACT preprints use data aggregated using the administrative regions of England, whereas for the purposes of this study the data has been aggregated using NHS regions. Additionally, small changes can occur in the aggregated datasets that were published in real time because of participant withdrawals and additional data cleaning.

##### 1.1.6 Summary of the data used for calibration

Table S 1 details the datasets used to calibrate the model to the regional epidemics.

**Table S 1:** Data sources and definitions.

| Data type | Description | Source | Reference |
| --- | --- | --- | --- |
| **Hospital deaths** | Daily number of COVID-19 deaths reported by NHS England within 28 days of a positive result | PHE | See data supplement. These data underlie what is released on (2) |
| **Care home deaths** | Daily number of COVID-19 deaths not reported by NHS England within 28 days of a positive result | PHE | See data supplement. These data underlie what is released on (2) |
| **ICU occupancy** | Daily number of confirmed COVID-19 patients in ICU | Gov.uk Dashboard | (2) |
| **General bed occupancy** | Daily number of confirmed COVID-19 patients in non-ICU beds | Gov.uk Dashboard | (2) |
| **Admissions** | Daily number of confirmed COVID-19 patients admitted to hospital | Gov.uk Dashboard | (2) |
| **Pillar 2 testing** | Daily number of positive and negative PCR test results | PHE | See data supplement. These data underlie what is released on (2) |
| **REACT-1 testing** | Daily number of positive and negative PCR test results | REACT | (3) |
| **Serology** | Serology survey conducted on blood donors aged 15-65 | PHE | See data supplement, these data are collected as part of (4) |
| **Patient progression in hospital** | Number of hospital admissions going down each treatment route (e.g. ICU, stepdown care) and length of stay in each ward. | CHESS | (5) |

##### 1.1.7 Other data sources

###### 1.1.7.1 Patient progression in hospital

The COVID-19 Hospitalisation in England Surveillance System (CHESS) data consists of a line list of daily individual patient-level data on COVID-19 infection in persons requiring hospitalisation, including demographic and clinical information on severity and outcomes. We use the individual dates of progression through hospital wards, from admission to eventual death or discharge, to produce age-stratified estimates of hospital progression parameters to be passed to the wider transmission model (see Section 1.9.2 and (supplementary data file: *support_progression.csv, support_severity.csv*).

###### 1.1.7.2 Demographic data

We use data from the Office for National Statistics (ONS (6)) to get the number of individuals in each of the 17 age-groups, i.e. 16 five-year age bands (0-4, 5-9, …, 75-79) and an 80+ group. We get the number of care-home beds in England from (7) giving us the number of care-home beds for each NHS regions. We then got an estimate of the total population of care-home residents in the UK from (8) that we scaled down to the England population size, combined with the estimate of the total number of beds in England, we derived a value of the total occupancy of care-homes of 74.2%. We assumed that the occupancy is the same in all the NHS regions. Care-home residents are subtracted from the 4 oldest age group (5% from age 65-69, 5% age 70-74, 15% age 75-79 and 75% age 80+ (9)). We then assume a 1:1 ratio of care-home residents to care-home workers and assume that the care-home workers population is homogeneously distributed among the 25-65 population in the region.

The contact matrix between the 17 age-groups is based on the POLYMOD contact survey. See parameterisation for more details (10).

#### 1.2 Evidence synthesis

Figure S 1 shows the functional relationships between data sources, modelled outputs and parameters in our study.

**Figure S 1:**
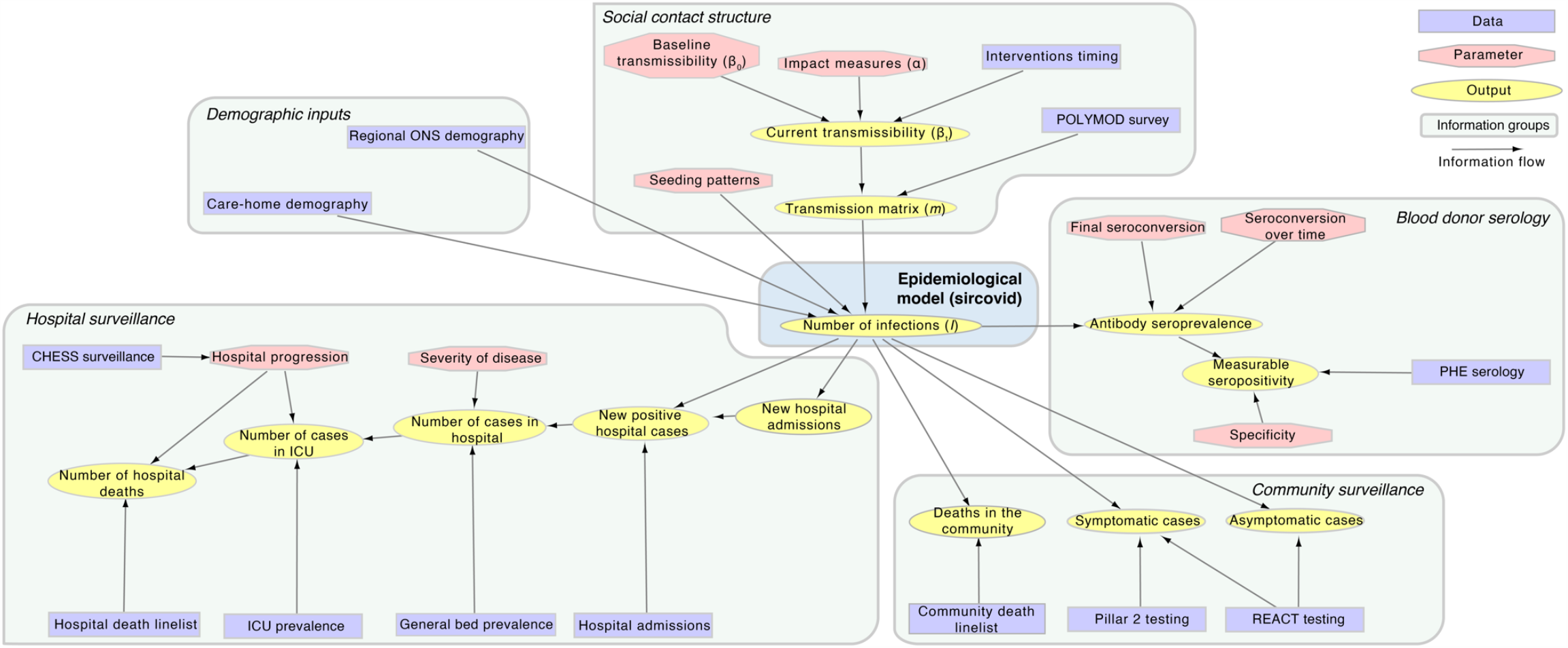
Graph showing the functional relationships between data sources (rectangles), modelled outputs (ovals) and parameters (hexagons).

#### 1.3 Model description

We developed a stochastic compartmental transmission-dynamic model incorporating hospital care pathways to reconstruct the course of the COVID-19 epidemic in the seven NHS regions of England (Figure S 2). All analyses were done by regions, and then aggregated somehow if needed (e.g. for national IFR, or cumulative incidence). In the following description we do not mention any index denoting the region and thus all notations refer to the same NHS region.

##### 1.3.1 Stratification of population into groups

We divided each regional population into 19 strata, denoted by the superscript *i*, 17 strata representing age groups within the general population, and two separate risk groups comprising care home workers (CHW) and care home residents (CHR). The 17 age groups consisted of 16 five-year age bands (0-4, 5-9, …, 75-79) and an 80+ group. The total size of the care home worker and resident groups were calculated assuming that 74.2% of available care home beds are occupied and there is a 1:1 carer to resident ratio (11). The care home workers were then split equally between all 8 age categories in the range 25 –64-year-old and removed from the corresponding age categories in the general population. Despite the care-home workers being removed from all age categories in the range 25 –64-year-old, they care-home workers are assumed to constitute one single group in our model for simplicity. The care home residents were drawn from the 65+ year old general population, such that 5% were aged 65-69, 5% aged 70-74, 15% aged 75-79 and 75% aged 80+ (9) and similarly removed from the corresponding age groups in the general population. Again, similarly to care-home workers they do constitute a single group in our model. We thus do not capture specific transmission dynamics within each care home, but rather an average mixing between residents and workers in the regional care home sector as a whole.

##### 1.3.2 Progression of infection and hospitalisation

Prior to the importation of COVID-19, all individuals were assumed equally susceptible to infection (*S*). Upon infection, individuals pass through a latent period (*E*) before becoming infectious. A proportion (*p*_*C*_) of infectious individuals develop symptoms (*I*_*C*_) while the rest remain asymptomatic (*I*_*A*_). All asymptomatic individuals are assumed to recover naturally. Those with symptoms may also recover naturally (*R*), however a proportion (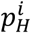, age/care home-dependant as indicated by the *i* superscript) develop severe disease requiring hospitalisation. Of these, a proportion 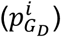 die at home without receiving hospital care. In practice this proportion is set to zero except among care home residents. Of the patients who are admitted to hospital, a proportion (*p*^∗^(*t*)) have their COVID-19 diagnoses confirmed prior to admission, while the remainder may be diagnosed during their inpatient stay. All hospital compartments are divided between suspected (but not yet confirmed) and confirmed diagnoses (indicated by superscript ^∗^). A proportion 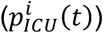 of new hospital admissions are triaged (*ICU*_*pre*_) before admission to the intensive care unit (*ICU*), where a fraction 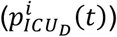 die; those who do not die get out of ICU to a ward (*W*) where a proportion 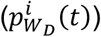 die, while the remainder recover, following an inpatient care stepdown period. Inpatients not triaged to the ICU are assigned to general hospital beds (*H*), where a proportion 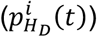 die, while the remainder recover. Recovered individuals are assumed to be immune to reinfection for at least the duration of the simulation.

In addition, there are two parallel flows which we use for fitting to testing data: (i) for PCR positivity and (ii) for seropositivity. Upon infection, an individual enters the PCR flow in a pre-positivity compartment 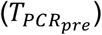 before moving into the PCR positivity compartment 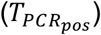 and then ultimately into the PCR negativity compartment 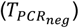. Meanwhile, individuals move into the seropositivity flow upon becoming infectious, entering first into a pre-seropositivity compartment 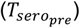. A proportion of individuals 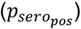 then seroconvert and move into the seropositivity compartment 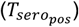, while the remainder move into the seronegativity compartment 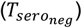.

We calibrated the duration distributions for each hospital compartment, and the age-stratified probabilities of moving between compartments, using the analysis of individual-level patient data (presented below in Section 1.9.2). The required Erlang distributional form was achieved within the constraints of the modelling framework by splitting each model compartment into *k* sequential sub-compartments (Table S 2).

**Figure S 2:**
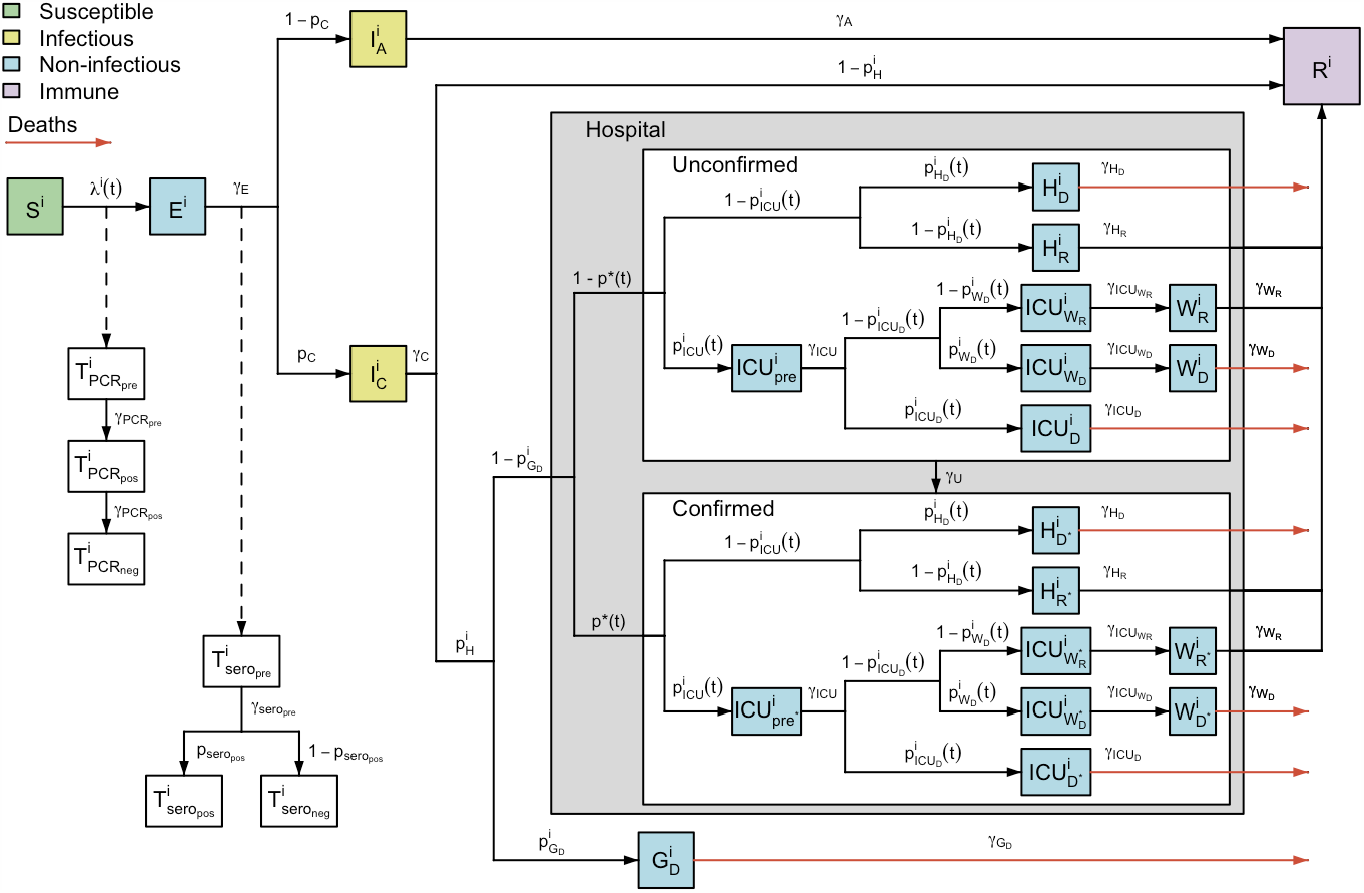
Model structure flow diagram with rates of transition between infection states. Variable names defined in text.

**Table S 2:** Description of model compartments and distribution of time spent in each. For each named compartment, we give the associated duration. Due to the Markovian structure these are model Erlang-like distributions with k_j_ the number of exponential-like compartments and γ_j_ the rate of the exponential-like compartment. 𝔼 [δ_j_] gives the mean duration in days spent in the corresponding compartment. The structure and duration of each stage was assumed to be the same for unconfirmed and confirmed cases in hospital (see Figure S2). For length of stays related to hospital pathways, more detail is given in section 1.9.2.

| Compartment | Description | Duration |  | (days) | Source |
| --- | --- | --- | --- | --- | --- |
| | | $\tau_j \sim \text{Erlang}(k_j, \gamma_j)$ | | $\mathbb{E}[\tau_j] = k_j/\gamma_j$ | |
| $j$ | | $k_j$ | $\gamma_j$ | (95% CI) | |
| $S$ | Susceptible to infection | Determined by transmission dynamics | | | |
| $E$ | Latent infection | 2 | 0.44 | 4.6 (0.6, 12.8) | Lauer et al.(12) |
| $I_A$ | Asymptomatic infection | 1 | 0.48 | 2.1 (0.1, 7.7) | Bi et al.(13) |
| $I_C$ | Symptomatic infection | 1 | 0.25 | 4.0 (0.1, 14.8) | Docherty et al.(14) |
| $G_D$ | Severe illness leading to death in the general population | 2 | 0.40 | 5.0 | Bernabeu-Wittel et al. (15) |
| $H_R$ | Hospitalised on general ward leading to recovery | 1 | 0.09 | 10.7 (0.3, 39.4) | Fitted to CHES |
| $H_D$ | Hospitalised on general ward leading to death | 2 | 0.19 | 10.3 (1.3, 28.8) | Fitted to CHES |
| $ICU_{pre}$ | Triage to ICU | 1 | 0.40 | 2.5 (0.1, 9.2) | Fitted to CHES |
| $ICU_{WR}$ | Hospitalised in ICU, leading to recovery | 1 | 0.06 | 15.6 (0.4, 57.6) | Fitted to CHES |
| $ICU_{WD}$ | Hospitalised in ICU, leading to death in step-down following ICU | 1 | 0.14 | 7.0 (0.2, 25.7) | Fitted to CHES |
| $ICU_D$ | Hospitalised in ICU, leading to death | 2 | 0.17 | 11.8 (1.4, 32.9) | Fitted to CHES |
| $W_R$ | Stepdown recovery period after leaving ICU | 2 | 0.16 | 12.2 (1.5, 34.0) | Fitted to CHES |
| $W_D$ | Stepdown period before death after leaving ICU | 1 | 0.12 | 8.1 (0.2, 29.7) | Fitted to CHES |
| $R$ | Recovered | - | - | - | - |
| $T_{pre}^{PCR}$ | Pre-PCR positive | 1 | 0.33 | 3.0 (0.1, 11.1) | Omar et al. (16) |
| $T_{pos}^{PCR}$ | True PCR positive | 1 | 0.06 | 17.5 (0.4, 64.5) | - |
| $T_{neg}^{PCR}$ | True PCR negative | - | - | - | - |
| $T_{pre}^{sero}$ | Pre-seroconversion | 1 | 0.08 | 13.0 (0.3, 48.0) | Benny et al. (17) |
| $T_{pos}^{sero}$ | True seropositive | - | - | - | - |
| $T_{neg}^{sero}$ | True seronegative | - | - | - | - |

Values of fitted parameters are set out in Table S 5.

##### 1.3.3 Progression of infection and hospitalisation

The force of infection, *λ*^*i*^(*t*), for individuals in group *i* ∈ {[0,5), …, [75,80), [80 +), *CHW, CHR*} depends on time-varying social mixing between age groups and prevalence in all age/care home groups:

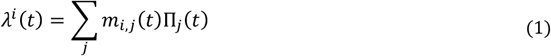

where *m*_*i,j*_ (*t*) is the (symmetric) time-varying person-to-person transmission rate from group j to group i, and Π_*j*_ (*t*) is the number of infectious individuals in group *j*, given by:

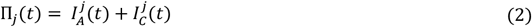

Broadly, to parameterise *m*_*i,j*_ (*t*), we informed mixing in the general population, and between the general population and care home workers using POLYMOD (10) via the R package *socialmixr* using age-structured regional demography (18).

Transmission between different age groups (*i,j*) ∈ {[0,5), …, [75,80), [80 +)}^2^ was parameterised as follows:

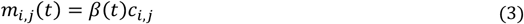

Here *c*_*i,j*_ is the (symmetric) person-to-person contact rate between age group i and j, derived from pre-pandemic data (10). *β* (*t*) is the time-varying transmission rate Dhich encompasses both changes over time in transmission efficiency (e.g. due to temperature) and temporal changes in the overall level of contacts in the population (due to changes in policy and behaviours).

We assumed *β* (*t*) to be piecewise linear:

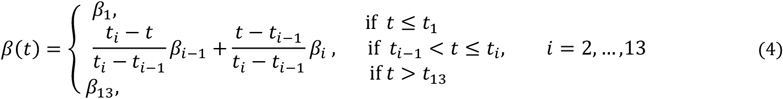

with 12 change points ***t*** _i_ corresponding to major announcements and changes in COVID-19 related policy, as detailed in Table S 3.

**Table S 3:** Changepoints for β (t)

| Changepoint | Value of $\beta(t)$ at changepoint | Date | Description |
| --- | --- | --- | --- |
| $t_1$ | $\beta_1$ | 16/03/20 | PM makes speech advising working from home, against non-essential travel (19) |
| $t_2$ | $\beta_2$ | 23/03/20 | PM announces lockdown 1 (20) |
| $t_3$ | $\beta_3$ | 25/03/20 | Lockdown 1 into full effect (21) |
| $t_4$ | $\beta_4$ | 11/05/20 | Initial easing of lockdown 1 (22) |
| $t_5$ | $\beta_5$ | 15/06/20 | Non-essential shops can re-open (23) |
| $t_6$ | $\beta_6$ | 04/07/20 | Restaurants, pubs etc can re-open (24) |
| $t_7$ | $\beta_7$ | 03/08/20 | "Eat out to help out" scheme starts (25) |
| $t_8$ | $\beta_8$ | 01/09/20 | Schools and universities re-open (26) |
| $t_9$ | $\beta_9$ | 14/09/20 | "Rule of six" introduced (27) |
| $t_{10}$ | $\beta_{10}$ | 14/10/20 | Tiered system introduced (28) |
| $t_{11}$ | $\beta_{11}$ | 31/10/20 | Lockdown 2 announced (29) |
| $t_{12}$ | $\beta_{12}$ | 05/11/20 | Lockdown 2 starts (29) |

The contact matrix *c*_*i,j*_ between different age groups (*i,j*) ∈ {[0,5), …, [75,80), [80 +)}^2^ is derived from the POLYMOD survey (10) for the United Kingdom using the *socialmixr* package (18,30), scaling by the local population demography to yield the required person-to-person daily contact rate matrix.

We defined parameters representing transmission rates within care homes (between and among workers and residents), which were assumed to be constant over time. Parameter *m*_*CHW*_ represents the person-to-person transmission rate among care home workers and between care home workers and residents; *m*_*CHR*_ represents the person-to-person transmission rate among care home residents. Hence,

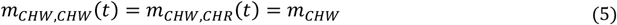

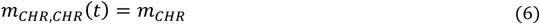

Transmission between the general population and care home workers was assumed to be similar to that within the general population, accounting for the average age of care home workers, with, for *i* ∈ {[0,5), …, [75,80), [80 +)},

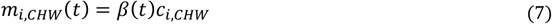

where *c*_*i,CHW*_ is the mean of *c*_*i*,[25,30)_, *c*_*i*,[30,35)_, …, *c*_*i*,[60,65)_ (i.e. of the age groups that the care home workers are drawn from).

Transmission between the general population and care home residents was assumed to be similar to that between the general population and the 80+ age group, adjusted by a reduction factor (∈, which was estimated), such that, for *i* ∈ {[0,5), …, [75,80), [80 +)},

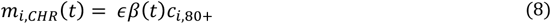

These represent contact between visitors from the general community and care home residents. This might involve a slightly different age profile than the age profile of the contact made by people in the 80+ age group.

##### 1.3.4 Age-varying and time-varying infection progression probabilities

Various probabilities of clinical progression within the model are assumed to vary across age groups to account for severity of infection varying with age, and some are assumed to vary in time in order to model improvements in clinical outcomes, such as those achieved through the use of dexamethasone (31).

Two probabilities are age-varying but not time-varying, the probability of admission to hospital for symptomatic cases, and the probability of death for severe symptomatic cases in care homes. These were modelled as follows:

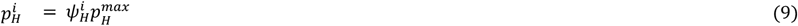

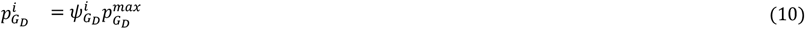

where for probability 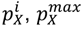 is the maximum across all groups and 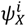 is the age scaling such that 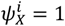 for the group corresponding to the maximum, against which all other groups are scaled.

As well as varying with age, four probabilities also vary with time: the probability of admission to ICU for hospitalised cases, the probability of death in ICU, the probability of death for hospitalised cases not admitted to ICU, and the probability of death in hospital after discharge from ICU:

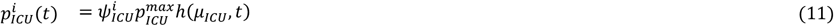

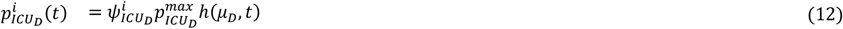

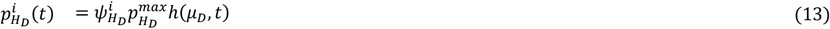

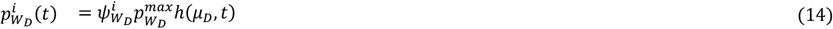

where here for probability 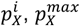 gives the maximum *initial* value across groups and *h*(*μ, t*) = 1 before April 1^st^, *h*(*μ*,, *t*) = *μ* < 1 after June 1^st^, with a linear reduction in between.

Care home residents with severe disease leading to death are assumed to remain in compartment **G**_**D**_ for 5 days on average before dying (modelled with 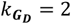 and 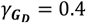), 95% range 0.6-13.9 days broadly consistent with durations in (15) and with duration about half the length observed in hospital streams (see Figure S *5*).

For care home workers, the age scaling 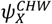 is taken as the mean of the age scalings 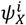 for *i* ∈ {[25,30), [30,35), …, [60,65)}. For care home residents, we assume that 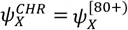, with the exception of the probability of individual with severe disease requiring hospitalisation dying at home (without receiving hospital care), where we assume 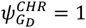 and 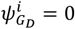 for all other groups, to effectively allow death outside hospital only for care home residents.

###### Reproduction number Y_W_ and effective reproduction number 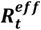

We calculated the reproduction number over time, *R*_*t*_, and effective reproduction number over time, 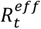, using next generation matrix methods (32). The reproduction numbers are calculated for the general population, i.e. excluding care home workers and residents. We define *R*_*t*_ as the average number of secondary infections a case infected at time t would generate in a large entirely susceptible population, and 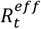 as the average number of secondary infections generated by a case infected at time t would accounting for the finite population size and potential immunity in the population.

To compute the next generation matrix, we calculated the mean duration of infectiousness Δ_*I*_, as

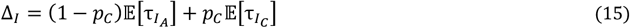

where parameter and model compartment notations are defined in Table S 2 - Table S 8. For this calculation, the expected durations of stay in compartments were adjusted to account for the discrete-time nature of the model, via calculating the expected number of time-steps (of length dt) spent in a given compartment. Note that if in continuous-time a compartment duration is *τ* ∼Erlang (*k, λ*), then the corresponding discrete-time mean duration is:

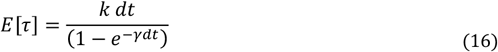

The next generation matrix was calculated as, for (i, j) ∈ {[0,5), …, [75,80), [80 +)}^2^,

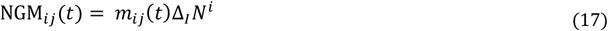

where N^i^ is the total population of group *i* and *R*_*t*_ is taken to be the dominant eigenvalue of NGM(*t*), while the effective next generation matrix was calculated as:

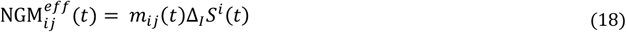

with 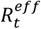 taken to be the dominant eigenvalue of NGM^eff^(*t*).

#### 1.5 Infection severity

Posterior estimates of severity, namely the infection hospitalisation and infection fatality ratios, were calculated in each group *i* as follows:

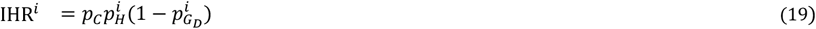

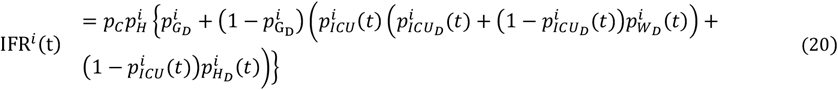

Note that for simplicity the notation we use do refer explicitly to the NHS region of interest. We calculated age-aggregated estimates for each region by weighting the age-specific severity estimates by the cumulative incidence in that age group. Aggregate estimates for England were then calculated by weighting the region-specific estimates by the regional attack rates.

#### 1.6 Compartmental model equations

To clearly illustrate the model dynamics, we describe a deterministic version of the model in differential equations (22)-(57), followed by the stochastic implementation used in the analysis. Each compartment is stratified by mixing category *i* ∈ {[0,5), …, [75,80), [80 +), CHW, CHR}. Full definitions of compartments and model parameters are set out in Table S 2 - Table S 8.

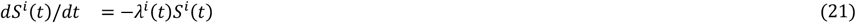

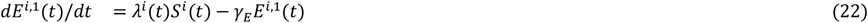

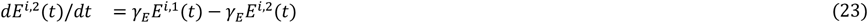

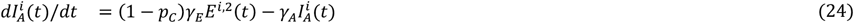

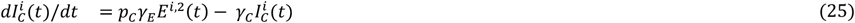

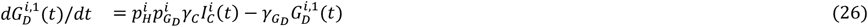

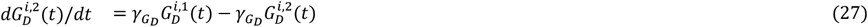

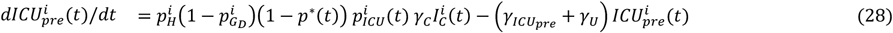

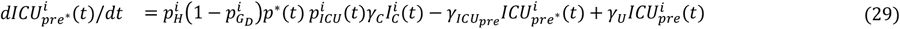

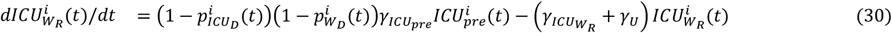

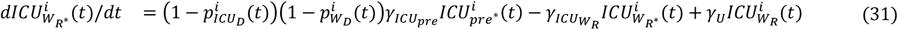

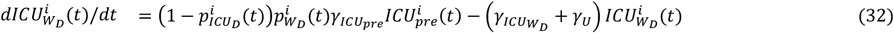

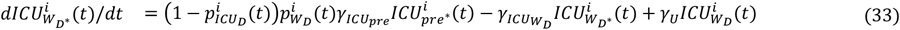

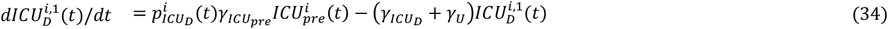

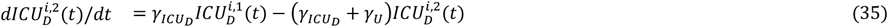

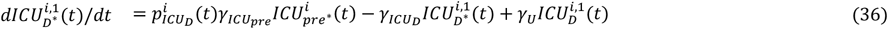

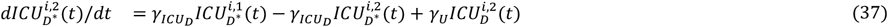

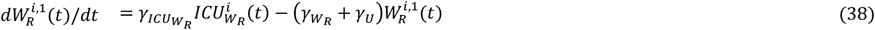

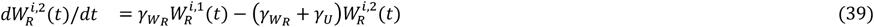

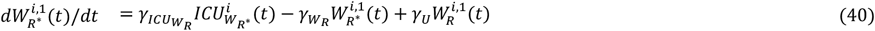

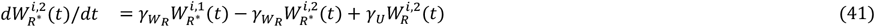

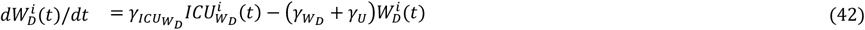

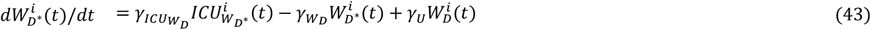

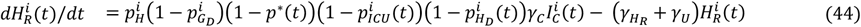

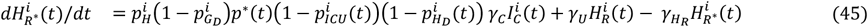

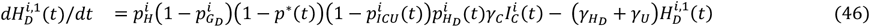

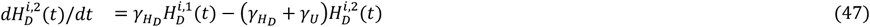

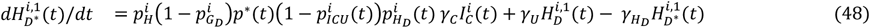

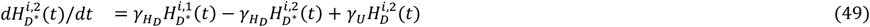

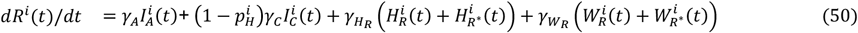

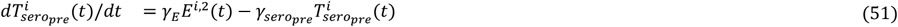

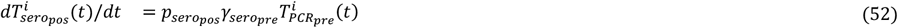

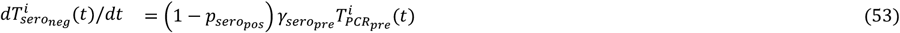

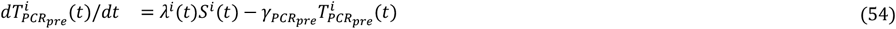

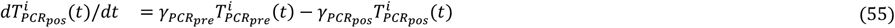

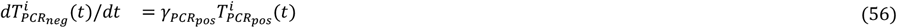

We used the tau-leap method (33) to create a stochastic, time-discretised version of the model described in equations (58-162), taking four update steps per day. The process was initialised with ten asymptomatic infectious individuals aged 15-19 on the epidemic start date *t*_0_, a parameter we estimate. For each time step, the model iterated through the procedure described below. In the following, we introduce a small abuse of notation: for transitions involving multiple onward compartments (e.g transition from compartment *E* to compartments *I*_*A*_ or *I*_*C*_), for conciseness, we write

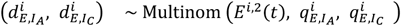

instead of

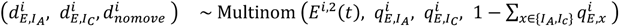

where 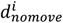 is a dummy variable counting the number of individuals remaining in compartment *E* ^*i*,2^.

Using this convention, transition variables are drawn from the following distributions, with probabilities defined below:

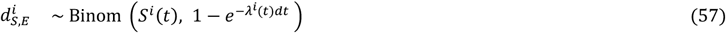

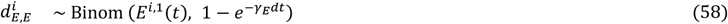

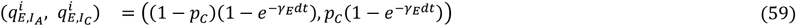

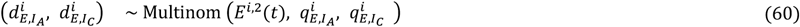

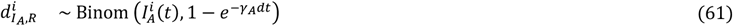

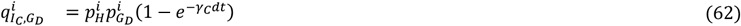

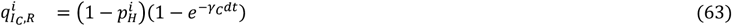

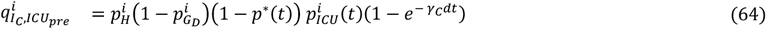

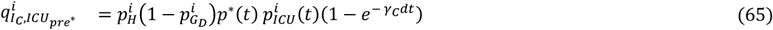

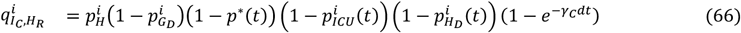

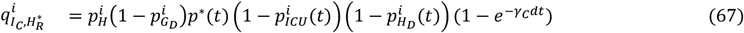

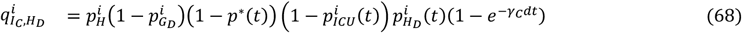

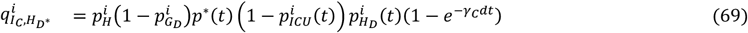

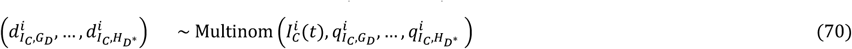

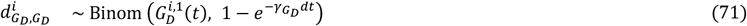

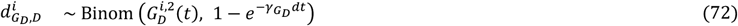

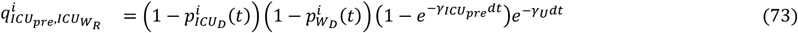

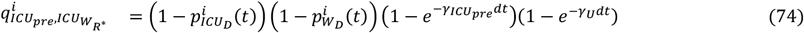

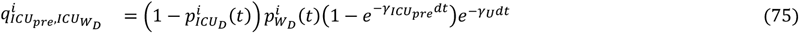

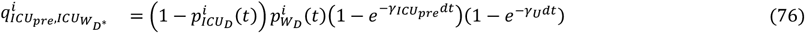

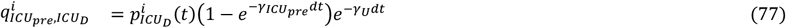

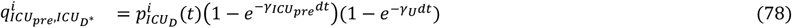

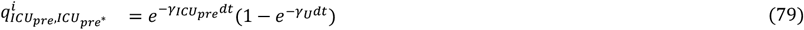

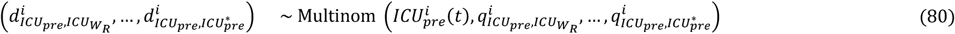

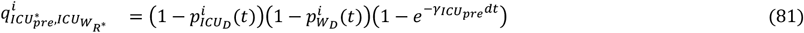

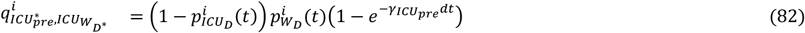

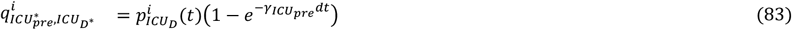

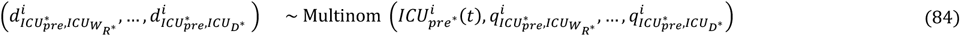

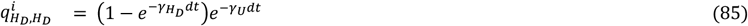

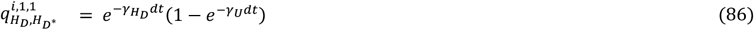

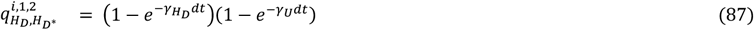

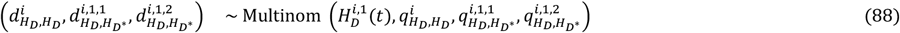

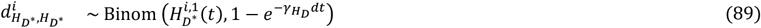

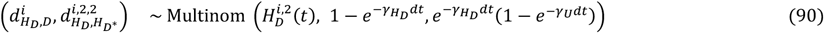

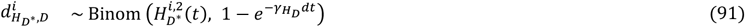

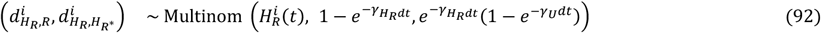

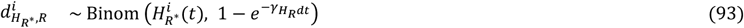

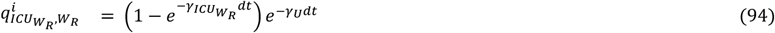

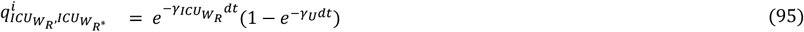

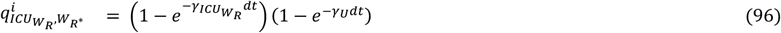

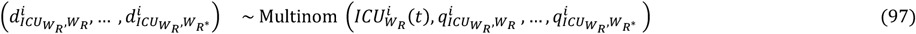

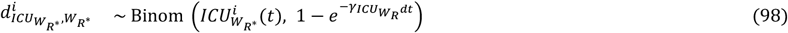

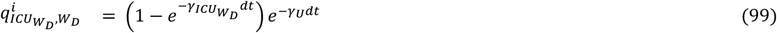

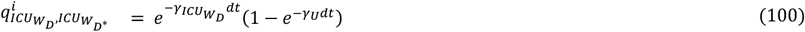

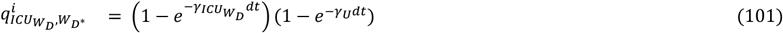

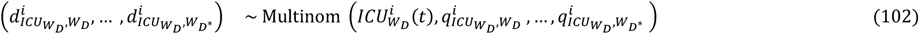

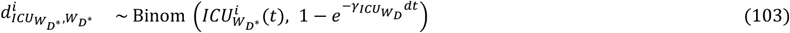

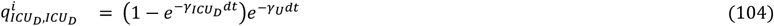

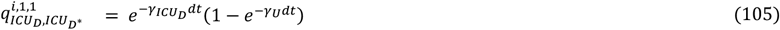

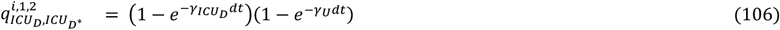

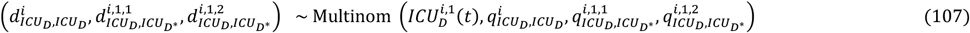

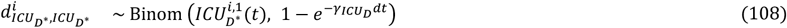

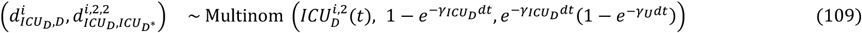

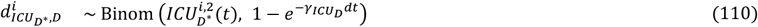

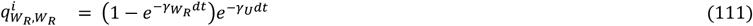

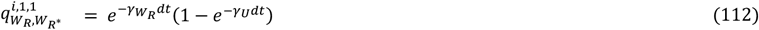

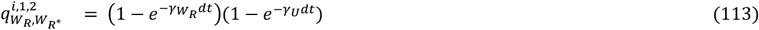

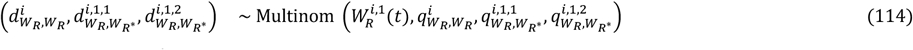

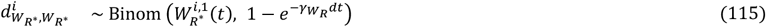

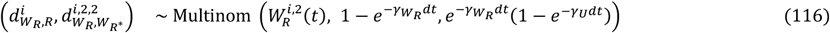

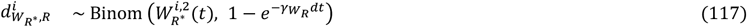

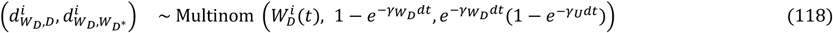

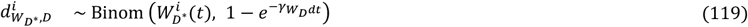

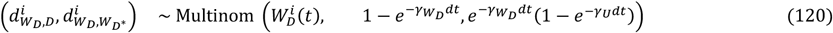

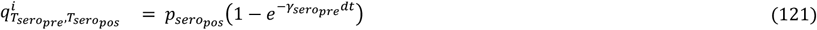

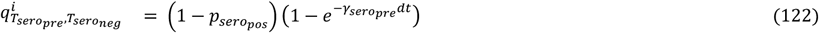

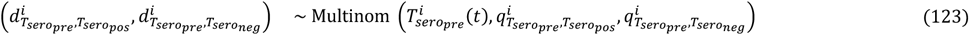

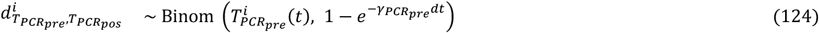

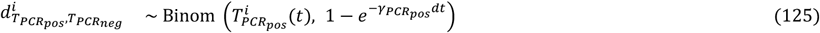

Model compartments were then updated as follows:

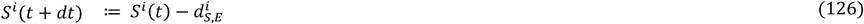

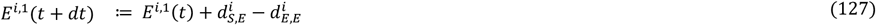

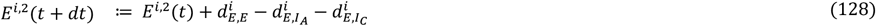

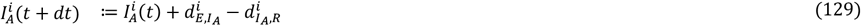

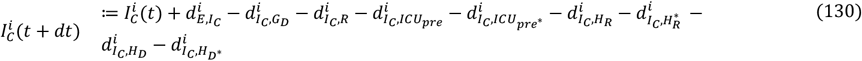

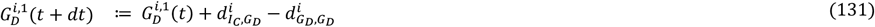

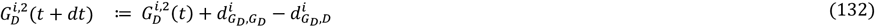

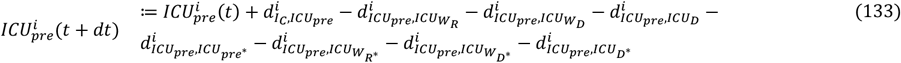

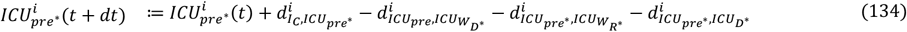

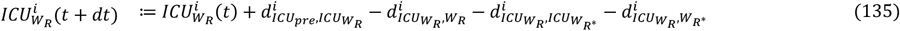

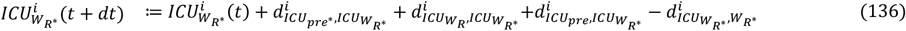

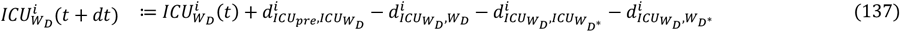

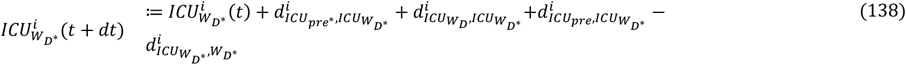

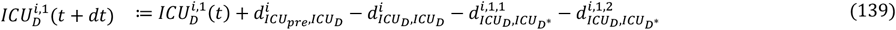

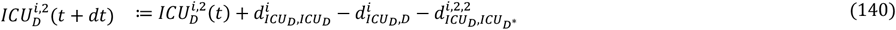

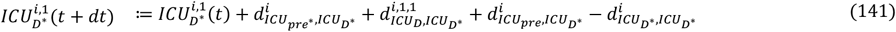

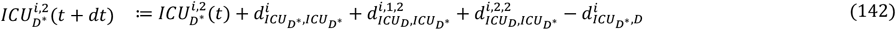

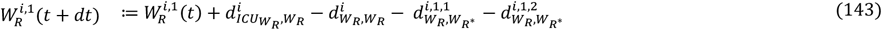

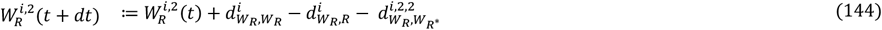

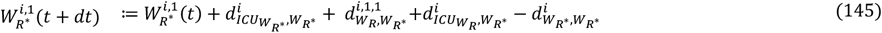

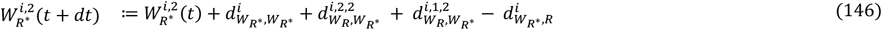

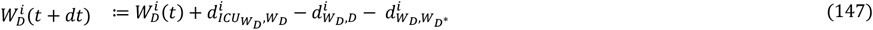

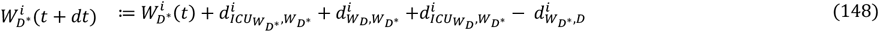

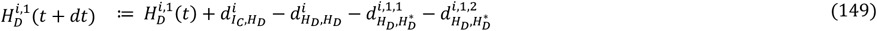

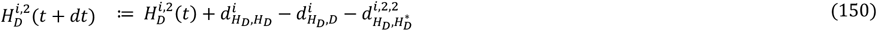

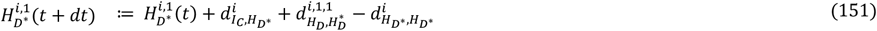

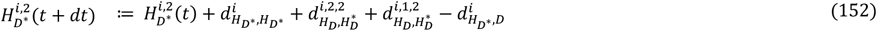

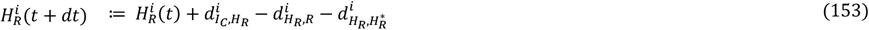

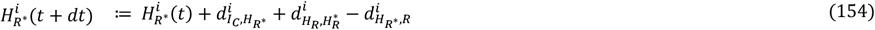

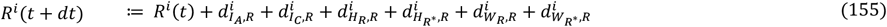

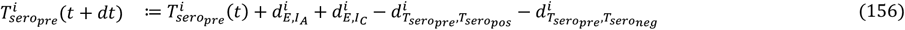

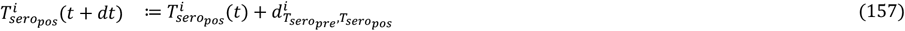

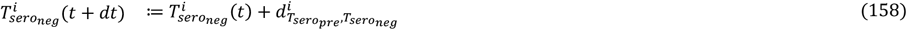

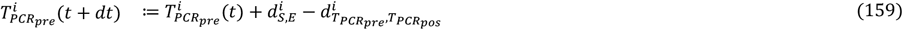

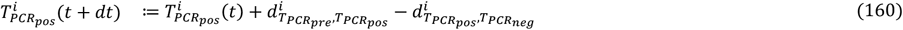

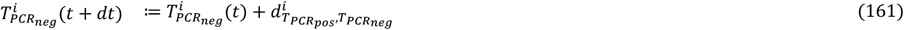

#### 1.7 Observation process

To describe the epidemic in each NHS region, we fitted our model to time series data on hospital admissions, hospital ward occupancy (both in general beds and in ICU beds), deaths in hospitals, deaths in care homes, population serological surveys and PCR testing data (section 1.1 and Table S 1).

##### 1.7.1 Notation for distributions used in this section

If *Y* ∼ NegBinom(*m, k*), then *Y* follows a negative binomial distribution with mean *m* and shape *κ*, such that

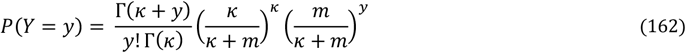

where Γ(*x*) is the gamma function. The variance of *Y* is 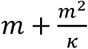.

If Z ∼ BetaBinom(*n, ω, ρ*), then *Z* follows a beta-binomial distribution with size *n*, mean probability ω and overdispersion parameter *ρ*, such that

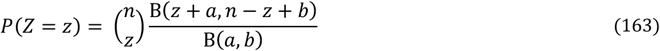

where 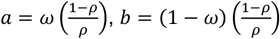 and B(*a, b*) is the beta function. The mean of Z is *nω* and the variance is *nω* (1 − *ω*)[1 + (*n* − 1) *ρ*].

##### 1.7.2 Hospital admissions and new diagnoses in hospital

We represented the daily number of confirmed COVID-19 hospital admissions and new diagnoses for existing hospitalised cases, *Y*_*adm*_ (*t*), as the observed realisations of an underlying hidden Markov process, *X*_*adm*_ (*t*), defined as:

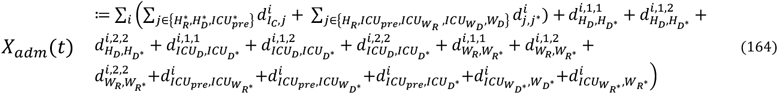

Which was related to the data via a reporting distribution:

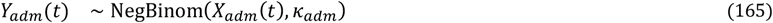

We allow for overdispersion in the observation process to account for noise in the underlying data streams, for example due to day-of-week effects on data collection. We adopt k = 2 for all NHSE data streams, so that they contribute equal weight to the overall likelihood.

##### 1.7.3 Hospital bed occupancy by confirmed COVID-19 cases

The model predicted general hospital bed occupancy by confirmed COVID-19 cases, *X*_*hosp*_ (*t*) as:

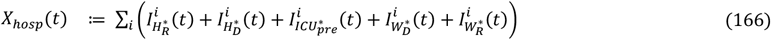

Which was related to the observed daily general bed-occupancy via a reporting distribution:

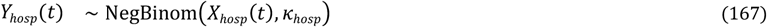

with *κ*_*hosp*_ = 2 as above.

Similarly, the model predicted ICU bed occupancy by confirmed COVID-19 cases, *X*_*ICU*_ (*t*) as:

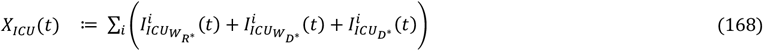

Which was related to the observed daily ICU bed-occupancy via a reporting distribution:

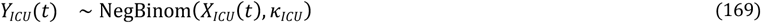

with *κ*_*ICU*_ = 2.

##### 1.7.4 Hospital and care homes COVID-19 deaths

The reported number of daily COVID-19 deaths in hospitals, 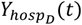 was considered as the observed realisation of an underlying hidden Markov process, 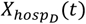, defined as:

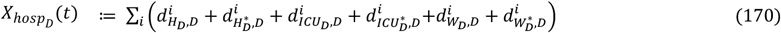

Which was related to the data via a reporting distribution:

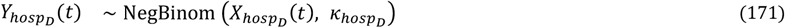

with 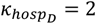.

Similarly, we represented the reported number of daily COVID-19 deaths in care homes, 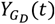, as the observed realisations of an underlying hidden Markov process, 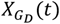, defined as:

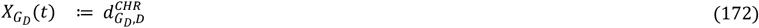

Which was related to the data via a reporting distribution:

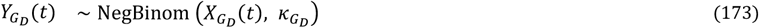

with 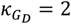.

##### 1.7.5 Serosurveys

We model serological testing of all individuals aged 15-65, and define the resulting number of seropositive and seronegative individuals (were all individuals aged 15-65 to be tested), as:

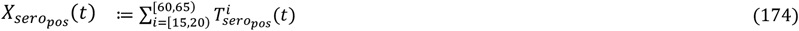

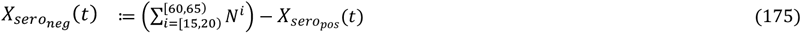

We compared the observed number of seropositive results, 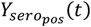, with that predicted by our model, allowing for i) the sample size of each serological survey, 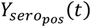and ii) imperfect sensitivity 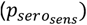 and specificity 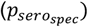 of the serological assay:

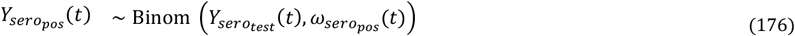

Where:

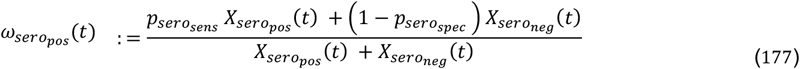

#### 1.7.6 PCR testing

As described in the data section (1.1), we fitted the model to PCR testing data from two separate sources:

- pillar 2: the government testing programme, which recommends that individuals with COVID-19 symptoms are tested (34),
- the REACT-1 study, which aims to quantify the prevalence of SARS-CoV-2 in a random sample of the England population on an ongoing basis (35).

We only use Pillar 2 PCR test results for individuals aged 25 and over (we assume this includes all care home workers and residents). We assume that individuals who get tested through Pillar 2 PCR testing are either newly symptomatic SARS-CoV-2 cases (who will test positive):

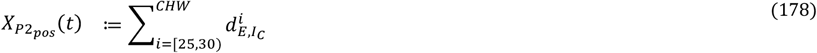

or non-SARS-CoV-2 cases who have symptoms consistent with COVID-19 (who will test negative):

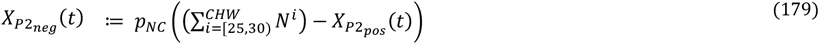

where *p*_*NC*_ is the probability of non SARS-CoV-2 cases having symptoms consistent with COVID-19 leading them to seek a PCR test.

We compared the observed number of positive PCR tests, 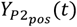 with that predicted by our model, accounting for the number of PCR tests conducted each day under pillar 2, 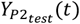, by calculating the probability of a positive PCR result (assuming perfect sensitivity and specificity of the PCR test):

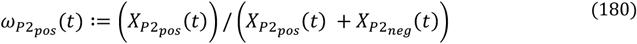

People may seek PCR tests for many reasons and thus the pillar 2 data are subject to competing biases. We therefore allowed for an over-dispersion parameter 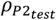, which we fitted separately for each region in the modelling framework:

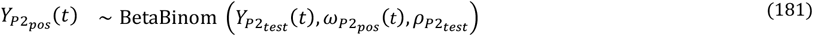

We incorporated the REACT-1 PCR testing data into the likelihood analogously to the serology data, by considering the model-predicted number of PCR-positives 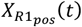 and PCR-negatives 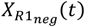, were all individuals aged over five and not resident in a care home to be tested:

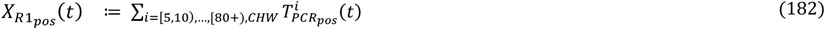

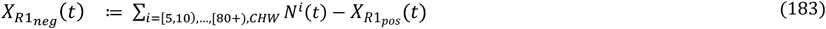

We compared the daily number of positive results observed in REACT-1, 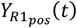, given the number of people tested on that day, 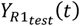, to our model predictions, by calculating the probability of a positive result, assuming perfect sensitivity and specificity of the REACT-1 assay:

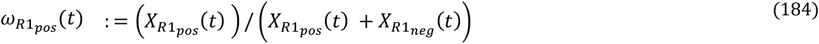

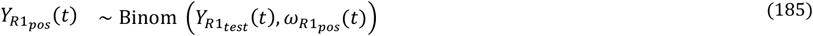

The overall likelihood function was then calculated as the product of the likelihoods of the individual observations.

#### 1.8 Bayesian inference and model fitting

A closed-form expression of the likelihood of the observed data given the model and its parameters was not analytically tractable, so we used particle filtering methods to obtain an unbiased estimate of the likelihood which can be efficiently sampled from (36). Where appropriate, we used estimates from the literature to set model parameters at fixed values. We limited the parameters being inferred to just those with particular epidemiological interest, or with large uncertainty in existing literature.

The model was fitted independently to each NHS region. For each NHS region, we aimed to infer the values of 26 model parameters:

- the local epidemic start-date, *t*_0_;
- thirteen transmission rates at different time points *β*_1_, …, *β*_12_;
- three parameters governing transmission to and within care homes m mCHW, mCHR, *ϵ*;
- the probability of symptomatic individuals developing serious disease requiring hospitalisation, 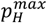, for the group with the largest probability;
- the probability of a care home resident dying in a care home if they have severe disease requiring hospitalisation, 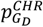;
- the probability, at the start of the pandemic, of a patient being admitted to ICU after hospitalisation, 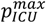, for the group with the largest probability;
- the probabilities, at the start of the pandemic, of dying in a hospital general ward, 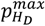, in the ICU, 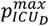, and in a stepdown ward following ICU, 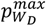, for the groups with the largest probability;
- the multiplier for hospital mortality after improvement in care, *μ*_H_;
- the multiplier for probability of admission to ICU after improvement in care, *μ*_*ICU*_;
- the daily proportion *p*_NC_, of the population seeking to get tested for an infection of SARS-Cov-2 following COVID-19 like symptoms and the overdispersion of the corresponding observation distribution 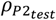.

We used particle Monte Carlo Markov Chain (pMCMC) methods (37), implementing a particle marginal Metropolis-Hastings algorithm with a bootstrap particle filter (38) with 96 particles (for sufficient variance in likelihood and a convenient multiple of number of available CPU cores for efficiency), to obtain a sample from the posterior distribution of the model parameters given the observed data. If the expected values of count distributions are zero when observed values are non-zero, this results in particles of zero weight, which can lead to the particle filter estimating the marginal likelihood to be 0. Therefore, to get a small but non-zero weight for each particle at every observation, within our particle filter likelihood we add a small amount of noise (exponentially distributed with mean 10^−6^) to count values from the model.

Within our particle filter we add small amounts of exponentially-distributed noise (with mean 10^−6^) to model outputs prior to calculating likelihood weights to avoid particles of zero weight, instead resulting in small but non-zero weights.

We implemented our model and parameter inference in an R package, *sircovid* (39), available at https://mrc-ide.github.io/sircovid, which uses two further R packages, *dust* to run the model in efficient compiled code and *mcstate* to implement the pMCMC sampler using Metropolis-Hastings sampling (40).

At each iteration, the sampler proposes an update to the joint distribution of parameters. These proposals are generated from multivariate Gaussian densities centred on the current parameter values, and with a covariance structure chosen to facilitate efficient mixing of the Markov chain. We specified reflecting boundaries for the proposal kernel to ensure that the proposed parameters are both epidemiologically and mathematically plausible and retain symmetry in the proposals.

For each regional fit, eight parallel chains of the pMCMC were run for 11,000 iterations, with the first 1,000 discarded as burn-in, and a 1/80 thinning. We assessed convergence visually.

#### 1.9 Prior distributions and parameter calibration

##### 1.9.1 Risk of hospital admission

In our Bayesian inference framework, we estimate 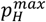, the probability of hospital admission for symptomatic cases in the group (across all ages and CHW and CHR) with the largest probability of hospital admission. However, we fix the relative probability of hospital admission for the other age groups, 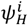, defined so that 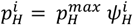, with 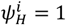 in the group with largest probability of hospital admission.

In this section we explain how the values of 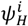 were chosen. We used two sources of information, an individual-level and an aggregated dataset. On the one hand, the COVID-19 Hospitalisation in England Surveillance System (CHESS) is a daily, confidential line list containing highly detailed information on patients admitted to hospital with confirmed COVID-19 (see following section 1.9.2 for further details). On the other hand, the Government’s Coronavirus Dashboard is an aggregated, publicly available situation report updated daily. Amongst other data, it provides updates on the number of daily admissions and hospital occupancy by devolved nation and, for England, by NHS region. We found the demography of hospitalisation in CHESS to be biased toward older patients compared to Dashboard data (Figure S3). We thus undertook a two-step approach to infer the demographic composition of COVID-19 hospitalisations across England.

Firstly, we derived an initial approximation of 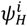 by dividing the total number of hospital admissions for age group *i* in CHESS over the total number of positive PCR tests (Pillar 2) for *i*. Both data sources were censored to include patients admitted to hospital or with a specimen data (i.e. the date the test was taken), respectively, between March 1 and December 2, 2020. We ran our full inference framework using this initial approximation for 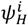 and observed its fit to the demographic composition of admissions from the data.

As a second step, we refined our initial approximations of 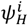 over a series of iterations of our inference framework, by drawing the modelled 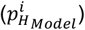 and observed 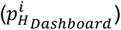 proportion of admissions for each age group (i.e. admissions in age group *i* divided by all admissions) and using it to derive a re-scaling factor for a new proposal for 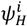 as follows:

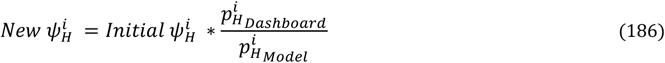

This process was repeated to obtain a close approximation to the observed proportion of admissions by age and region (Figure S3). A key strength of our approach is that we did not overfitted demography by individual regions. Rather, by assuming 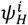 to be independent of geographic region, we allowed our inference framework to derive the number of admissions for each five-year age band *i* solely based on 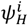, the demographic composition of the NHS region and inferred epidemic parameters, such as *R*_*t*_.

**Figure S 3:**
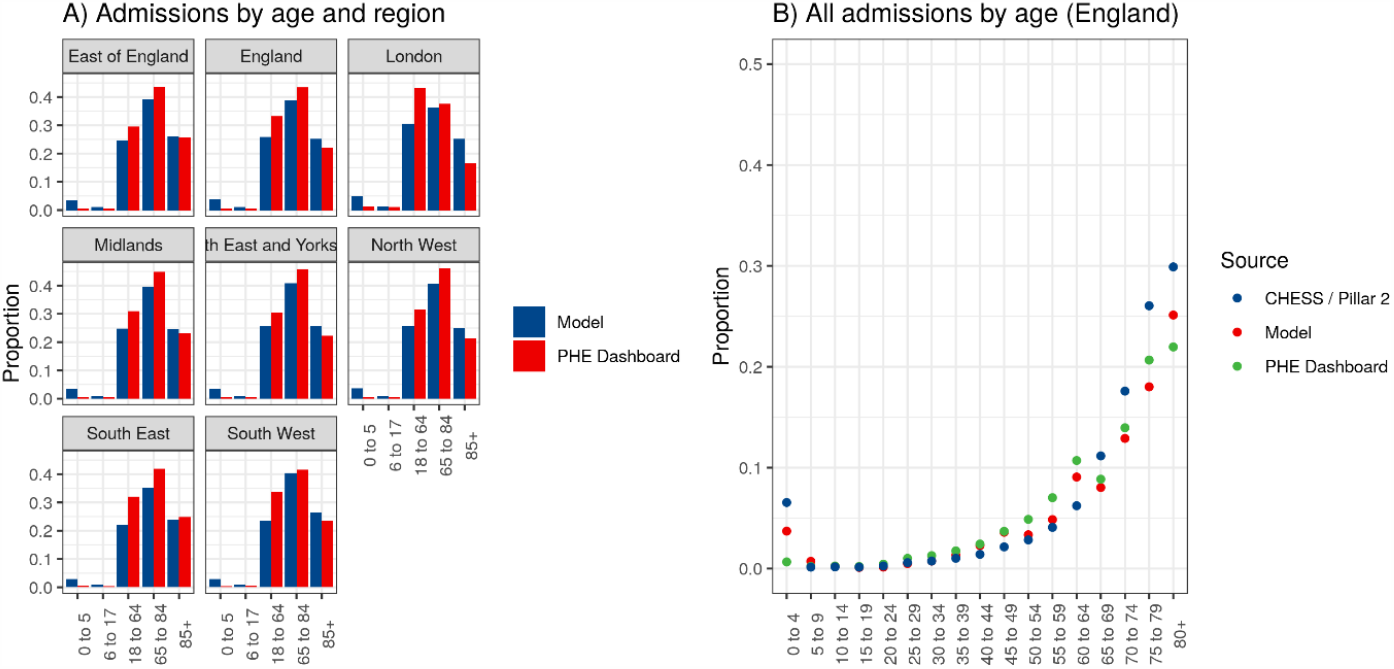
Proportion of admissions by age. a) Comparison of model outputs to data from the Government’s Coronavirus Dashboard, aggregated to five broad age categories. b) Age spline fitted (red) to Government’s Coronavirus Dashboard, with age categories disaggregated to five-year bands. The fitted spline (red) was used as input parameters for the probability of hospitalisation by age.

##### 1.9.2 Severity and hospital progression

We also performed extensive preliminary analysis to inform the age-structure of progression parameters within hospital. Data from the COVID-19 Hospitalisation in England Surveillance System (CHESS) were used to fit a simple model of patient clinical progression in hospital. The model structure was designed to mirror the within-hospital component of the wider mechanistic transmission model, but without the complexities arising from unknown admission dates and with greater detail on trends with age (Figure S 4).

**Figure S 4:**
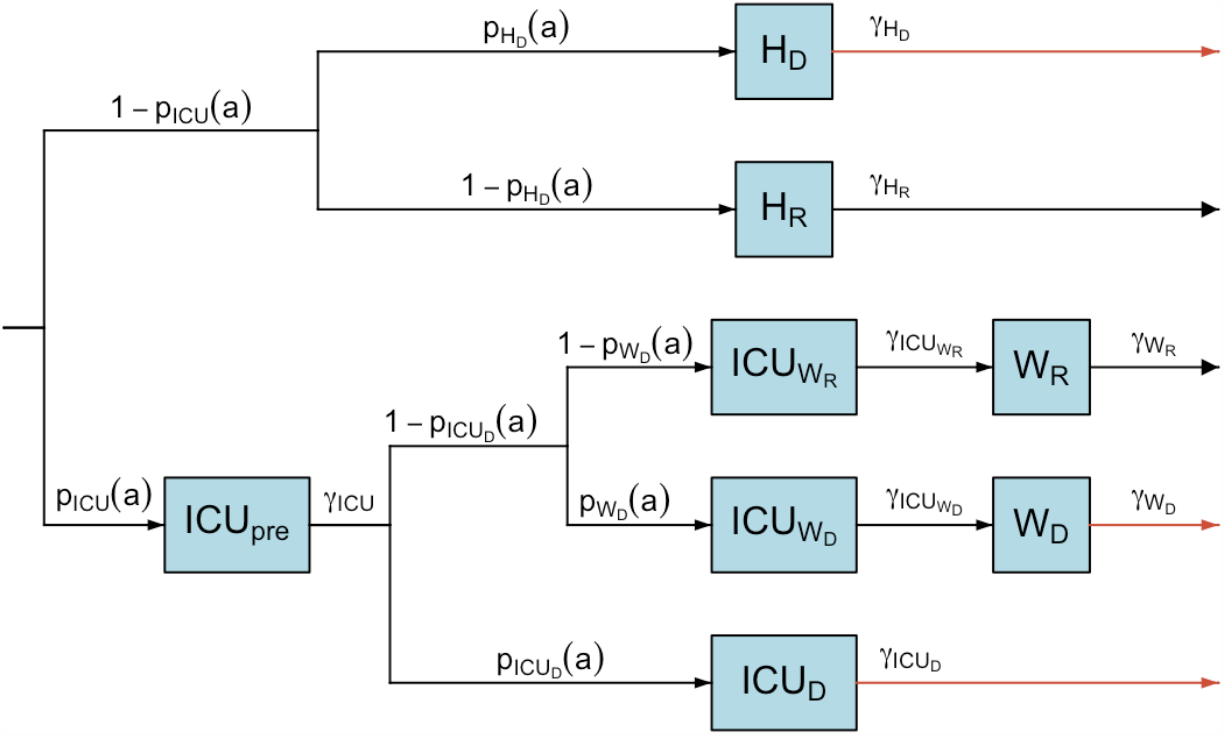
Directed Acyclic Graph of the hospital pathways fitted to CHESS data, which mirror the model structure described in Figure S 2, but with all parameters varying with age and not over time.

CHESS data consists of a line list of daily individual patient-level data on COVID-19 infection in persons requiring hospitalisation, including demographic and clinical information on severity and outcomes. Data were filtered to patients admitted between 18^th^ March and 31^st^ May 2020 (inclusive), with subsequent progression events possible up until 25^th^ Nov 2020.

This gave >5 months for outcomes to complete, and hence justified filtering to patients with resolved outcomes only. The length of stay in each state was taken as the difference between the registered dates of entering and leaving each hospital ward. Lengths of stay were assumed to follow Erlang distributions, as in the wider model, with a distinct mean and shape parameter for each state. Specifically, the probability of being in state 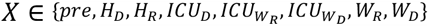 for *n* ∈ ℕ_0_ days was taken as the integral over day n of the Erlang distribution with mean *m*_*X*_ and shape *s*_*X*_:

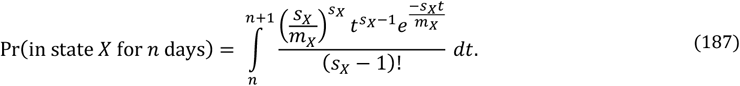

For a patient of age *a*, this was combined with the probability of their path through the hospital progression model, taken as the product of the individual transition probabilities at each bifurcation, i.e. values taken from *p*_*z*_(a) for *Z* ∈ {*ICU, H*_*D*_, *ICU*_*D*_, *W*_*D*_}. Transition probabilities were modelled as functions of age using logistic-transformed cubic splines. Knots were defined at coordinates 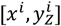, where *x*^*i*^ values were fixed at {0, 20, 40, 60, 80, 100, 120} and 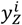 were free parameters to be estimated. The complete spline, *y*_*z*_(a) for a ∈ 0: 120, was obtained from these knots using standard expressions for cubic spline interpolation. Finally, transition probabilities were obtained from the raw *y*_*z*_(a) values using the logistic transformation:.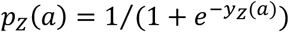.

In total there were 44 free parameters in the within-hospital progression model: 8 mean length of stay parameters, 8 length of stay shape parameters and 4 × 7 transition probability spline nodes (Figure S 4, Table S 4).

**Table S 4:** Descriptions of all states and transitions in the simplified hospital progression model fitted to CHESS data.

| State (X) | Description |
| --- | --- |
| $pre$ | General admission before step-up to ICU |
| $H_D$ | General ward before death in general ward |
| $H_R$ | General ward before discharge from general ward |
| $ICU_D$ | ICU before death in ICU |
| $ICU_{W_D}$ | ICU before step-down and eventual death in step-down care |
| $ICU_{W_R}$ | ICU before step-down and eventual discharge from step-down care |
| $W_D$ | Step-down (general) ward before death |
| $W_R$ | Step-down (general) ward before discharge |
| Transition (Z) | Description |
| $ICU$ | Admission to ICU from general ward |
| $H_D$ | Death in general ward |
| $ICU_D$ | Death in ICU |
| $W_D$ | Death in step-down care |

All parameters of the hospital progression model were given priors (Table S 5) and estimated within a Bayesian framework. All length of stay parameters were given uniform priors over a plausible range of values. For transition probabilities, the first spline node 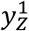 was given a prior that corresponded to a uniform distribution after logistic transformation, and subsequent spline nodes were given a multivariate normal prior to apply a smoothing constraint to the spline. Parameters were estimated jointly via MCMC using the custom package *markovid* v1.5.0 (41), which uses the random-walk Metropolis-Hastings algorithm to draw from the joint posterior distribution. MCMC was run for 1000 burn-in iterations and 100,000 sampling iterations replicated over 10 independent chains. Convergence was assessed via the Gelman-Rubin diagnostic (all parameters had potential scale reduction factor <1.1) and sampling sufficiency was assessed by visualising posterior distributions and by effective sample size (ESS) calculations (all parameters had ESS >100,000).

**Table S 5:** Priors on all length of stay distributions and transition probability splines. Uniform_cont_(a, b) denotes the continuous uniform distribution, and Uniform_disc_(a, b) the discrete uniform distribution between a and b (inclusive).

| Parameter | Description | Prior |
| --- | --- | --- |
| $m_X = \frac{1}{\gamma_X}$ | Mean of Erlang length of stay distribution | $m_X \sim Uniform_{cont}(0, 20)$ |
| $k_X$ | Shape parameter of Erlang length of stay distribution | $k_X \sim \text{Uniform}_{\text{disc}}(1,10)$ |
| $y_Z^1$ | First spline node of (transformed) transition probability | $f(y_Z^1) \propto \frac{e^{-y_Z^1}}{(1+e^{-y_Z^1})^2}$,<br>for $y_Z^1 \in (-10, 10)$ |
| $y_Z^j$ for $j \in 2:7$ | Subsequent spline nodes of (transformed) transition probability | $y_Z^j \sim \text{Normal}(y_Z^{j-1}, 0.25)$ |

Parameter estimates (posterior medians) were passed to the wider mechanistic transmission model as fixed values (Figure S *5*). For transition probabilities, the full age-spline (Figure 3, main text) was aggregated to 5-year age groups and normalised by the largest value to define the relative risk with age. The absolute risk in the mechanistic transmission model was obtained by multiplying the relative risk by region-specific scaling factors that were fitted as free parameters in the pMCMC. Hence, the preliminary analysis of CHESS data was used to inform trends of severity with age, but not the absolute probability of progression through the hospital states, which was informed by the Government’s Coronavirus Dashboard data.

For the wider mechanistic transmission model, we used Beta distributions for the priors of the various fitted probabilities regarding hospitalisation. The priors for 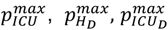 and 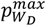 were all informed by the fitting to CHESS data by taking the median fitted value for the prior mean, which we halve in the case of 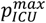 to account for CHESS being biased to more severe patients. The prior distributions are then calibrated so that the lower bound of the 95% confidence interval is 0.1 lower than the prior mean. For 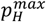 and 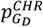, we assume prior means of 0.75 and calibrate the prior so that the lower bound of the 95% confidence interval is 0.2 lower than the mean. For the multipliers for hospital mortality after improvement in care, *μ*_*H*_, and for probability of admission to ICU after improvement in care, *μ*_*ICU*_ we used uninformative, [0,1] priors.

**Figure S 5:**
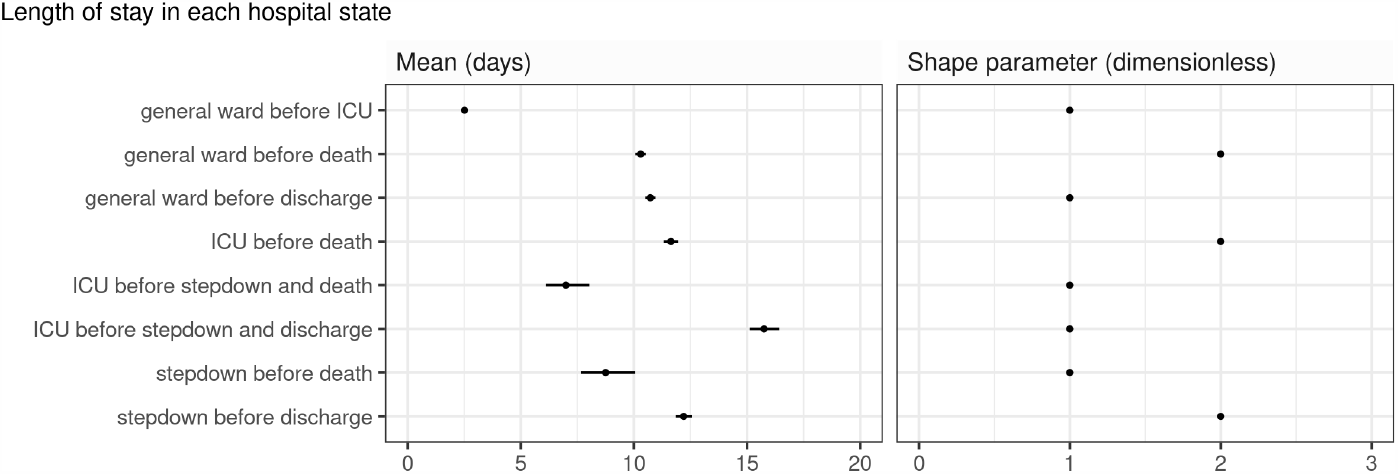
Posterior 95% credible intervals of length of stay mean (left) and shape parameters (right).

##### 1.9.3 Serosurveys

To keep serology parameters consistent between all regions we used estimates from the literature to fix the parameters of the seroconversion process. An alternative would have been to use these estimates as priors within a hierarchical model where some parameters would be shared between regions, but this would be much more involved computationally.

As described in section 1.3.2, the time to seroconversion from leaving the *E*^*i*^ compartment is modelled by an exponential distribution time spent in 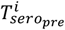 with a proportion 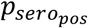 ultimately seroconverting and moving to 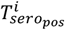 and the remaining staying negative and moving to 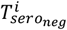.

We fixed 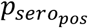 to 0.85 based on the estimate of 85% of infections becoming detectably seropositive with the EUROIMMUN assay used in the NHSBT serological surveys (42). The specificity of the serology test is 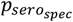 fixed to 0.99 also from (42). Finally, the sensitivity of serology test 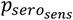 is assumed to be 1 as it is non-distinguishable from the time varying seroconversion process (Table S7).

##### 1.9.4 PCR positivity

As for other compartments, we modelled the duration of SARS-CoV-2 PCR-positivity after symptom onset using an Erlang distribution *τ* ∼Erlang(*k, γ*), with k successive compartments and a total mean time spent of 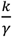 and variance 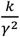.

We estimated the parameters of this distribution from Omar et al. (16), which reported the cumulative distribution of duration of PCR positivity in 523 individuals with mild COVID-19 disease in home quarantine in a German region. We performed a survival analysis using a gamma-accelerated failure time model fitted to their data, from which we estimated the mean and variance of the time from symptom onset to PCR negativity. This was used to derive values of k and *U* shown in Table S 2.

##### 1.9.5 Local start date of the epidemic

The start date of the epidemic for each region is assumed to have a uniform prior on the dates from 1^st^ January 2020 to 15^th^ March 2020, inclusive –with the latter date corresponding to the last date before the data begin.

##### 1.9.6 Time-varying transmission rates

We set priors for the transmission rates *β*_1_, …, *β*_12_ to reflect a Gamma distribution for the reproduction number *R*_*t*_ with a reasonable 95% confidence interval a priori. To obtain a prior for the corresponding *β*_*k*_, we then scale by a factor of 0.0241 (given other parameter values, *β*_*k*_ = 0.0241 would correspond approximately to *R*_*t*_ = 1). The 95% ranges for *R*_0_ we used are (i) (2.5, 3.5) at the onset of the epidemic (corresponding to *β*_1_); and then *R*_*t*_ (ii) (0.4, 3.5) at announcement of the first lockdown (corresponding to *β*_2_); and (iii) (0.4, 3) from the implementation of the first lockdown onwards (corresponding to *β*_3_, …, *β*_12_). The values are consistent with the values of the COMIX study (43).

##### 1.9.7 Transmission within care homes

For the transmission between care home workers and residents, *m*_*CHW*_, and transmission among care home residents, *m*_*CHR*_, we used a prior distributions reflecting that these are person-to-person infectious contact rates and thus should be scaled according to regional care home demography. We then used a Gamma distribution with shape 5 and mean 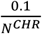 for both of these parameters (recall that we assume there is a 1-to-1 ratio of care home workers to residents in each region, so *N*^*CHW*^ = *N*^*CHR*^).

For the parameter governing the reduction in contacts between the general population and care home residents, *ϵ*, we used an uninformative *U*[0,1] prior.

##### 1.9.8 Parameters relating to Pillar 2 testing

For both the parameters *p*_*NC*_ and 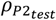, we used uninformative *U*[0,1] priors.

**Table S 6:**
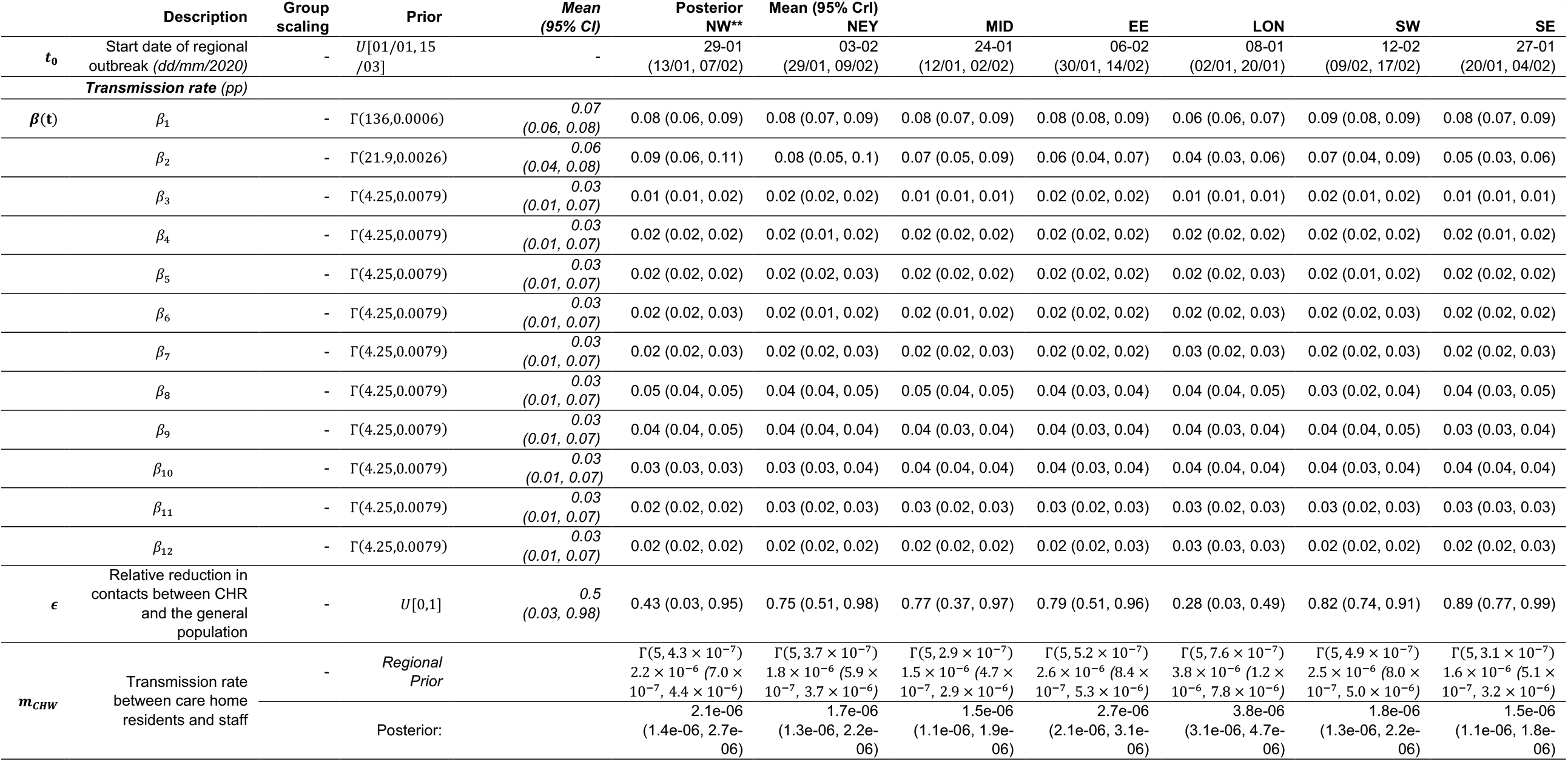
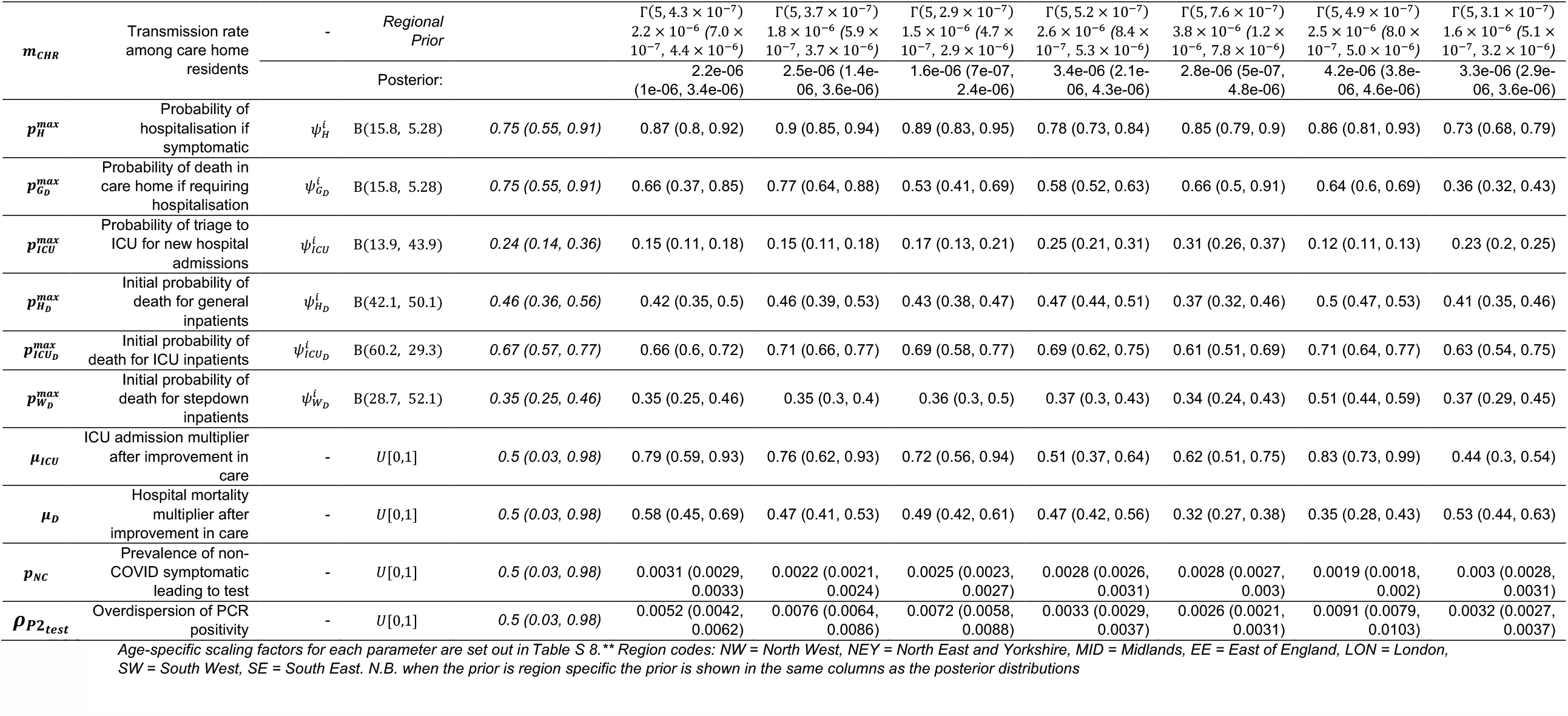
Inferred model parameter notations, prior and posterior distributions. Note that Γ (a, b) here refers to a Gamma distribution with shape a and scale b (such that the mean is ab), and ‘(a, b) refers to a Beta distribution with shape parameters a and b (such that the mean is a/(a + b)).

|  | Description | Group scaling | Prior | Mean (95% CrI) | Posterior NW** | Mean (95% CrI) NEY | MID | EE | LON | SW | SE |
| --- | --- | --- | --- | --- | --- | --- | --- | --- | --- | --- | --- |
| $t_0$ | Start date of regional outbreak (dd/mm/2020) | - | $U[01/01, 15/03]$ | - | 29-01 (13/01, 07/02) | 03-02 (29/01, 09/02) | 24-01 (12/01, 02/02) | 06-02 (30/01, 14/02) | 08-01 (02/01, 20/01) | 12-02 (09/02, 17/02) | 27-01 (20/01, 04/02) |
| **Transmission rate (pp)** |  |  |  |  |  |  |  |  |  |  |  |
| $\beta(t)$ | $\beta_1$ | - | $\Gamma(136, 0.0006)$ | $\frac{0.07}{(0.06, 0.08)}$ | 0.08 (0.06, 0.09) | 0.08 (0.07, 0.09) | 0.08 (0.07, 0.09) | 0.08 (0.08, 0.09) | 0.06 (0.06, 0.07) | 0.09 (0.08, 0.09) | 0.08 (0.07, 0.09) |
| | $\beta_2$ | - | $\Gamma(21.9, 0.0026)$ | $\frac{0.06}{(0.04, 0.08)}$ | 0.09 (0.06, 0.11) | 0.08 (0.05, 0.1) | 0.07 (0.05, 0.09) | 0.06 (0.04, 0.07) | 0.04 (0.03, 0.06) | 0.07 (0.04, 0.09) | 0.05 (0.03, 0.06) |
| | $\beta_3$ | - | $\Gamma(4.25, 0.0079)$ | $\frac{0.03}{(0.01, 0.07)}$ | 0.01 (0.01, 0.02) | 0.02 (0.02, 0.02) | 0.01 (0.01, 0.01) | 0.02 (0.02, 0.02) | 0.01 (0.01, 0.01) | 0.02 (0.01, 0.02) | 0.01 (0.01, 0.01) |
| | $\beta_4$ | - | $\Gamma(4.25, 0.0079)$ | $\frac{0.03}{(0.01, 0.07)}$ | 0.02 (0.02, 0.02) | 0.02 (0.01, 0.02) | 0.02 (0.02, 0.02) | 0.02 (0.02, 0.02) | 0.02 (0.02, 0.02) | 0.02 (0.02, 0.02) | 0.02 (0.01, 0.02) |
| | $\beta_5$ | - | $\Gamma(4.25, 0.0079)$ | $\frac{0.03}{(0.01, 0.07)}$ | 0.02 (0.02, 0.02) | 0.02 (0.02, 0.03) | 0.02 (0.02, 0.02) | 0.02 (0.02, 0.02) | 0.02 (0.02, 0.03) | 0.02 (0.01, 0.02) | 0.02 (0.02, 0.02) |
| | $\beta_6$ | - | $\Gamma(4.25, 0.0079)$ | $\frac{0.03}{(0.01, 0.07)}$ | 0.02 (0.02, 0.03) | 0.02 (0.01, 0.02) | 0.02 (0.01, 0.02) | 0.02 (0.02, 0.02) | 0.02 (0.02, 0.03) | 0.02 (0.02, 0.03) | 0.02 (0.02, 0.02) |
| | $\beta_7$ | - | $\Gamma(4.25, 0.0079)$ | $\frac{0.03}{(0.01, 0.07)}$ | 0.02 (0.02, 0.03) | 0.02 (0.02, 0.03) | 0.02 (0.02, 0.03) | 0.02 (0.02, 0.02) | 0.03 (0.02, 0.03) | 0.02 (0.02, 0.03) | 0.02 (0.02, 0.03) |
| | $\beta_8$ | - | $\Gamma(4.25, 0.0079)$ | $\frac{0.03}{(0.01, 0.07)}$ | 0.05 (0.04, 0.05) | 0.04 (0.04, 0.05) | 0.05 (0.04, 0.05) | 0.04 (0.03, 0.04) | 0.04 (0.04, 0.05) | 0.03 (0.02, 0.04) | 0.04 (0.03, 0.05) |
| | $\beta_9$ | - | $\Gamma(4.25, 0.0079)$ | $\frac{0.03}{(0.01, 0.07)}$ | 0.04 (0.04, 0.05) | 0.04 (0.04, 0.04) | 0.04 (0.03, 0.04) | 0.04 (0.03, 0.04) | 0.04 (0.03, 0.04) | 0.04 (0.04, 0.05) | 0.03 (0.03, 0.04) |
| | $\beta_{10}$ | - | $\Gamma(4.25, 0.0079)$ | $\frac{0.03}{(0.01, 0.07)}$ | 0.03 (0.03, 0.03) | 0.03 (0.03, 0.04) | 0.04 (0.04, 0.04) | 0.04 (0.03, 0.04) | 0.04 (0.04, 0.04) | 0.04 (0.03, 0.04) | 0.04 (0.04, 0.04) |
| | $\beta_{11}$ | - | $\Gamma(4.25, 0.0079)$ | $\frac{0.03}{(0.01, 0.07)}$ | 0.02 (0.02, 0.02) | 0.03 (0.02, 0.03) | 0.03 (0.02, 0.03) | 0.03 (0.02, 0.03) | 0.03 (0.02, 0.03) | 0.03 (0.02, 0.03) | 0.03 (0.03, 0.03) |
| | $\beta_{12}$ | - | $\Gamma(4.25, 0.0079)$ | $\frac{0.03}{(0.01, 0.07)}$ | 0.02 (0.02, 0.02) | 0.02 (0.02, 0.02) | 0.02 (0.02, 0.02) | 0.02 (0.02, 0.03) | 0.03 (0.03, 0.03) | 0.02 (0.02, 0.02) | 0.02 (0.02, 0.03) |
| $\epsilon$ | Relative reduction in contacts between CHR and the general population | - | $U[0,1]$ | $\frac{0.5}{(0.03, 0.98)}$ | 0.43 (0.03, 0.95) | 0.75 (0.51, 0.98) | 0.77 (0.37, 0.97) | 0.79 (0.51, 0.96) | 0.28 (0.03, 0.49) | 0.82 (0.74, 0.91) | 0.89 (0.77, 0.99) |
| $m_{CHW}$ | Transmission rate between care home residents and staff | - | Regional Prior | | $\Gamma(5, 4.3 \times 10^{-7})$<br>$2.2 \times 10^{-6}$ ($7.0 \times 10^{-7}$, $4.4 \times 10^{-6}$) | $\Gamma(5, 3.7 \times 10^{-7})$<br>$1.8 \times 10^{-6}$ ($5.9 \times 10^{-7}$, $3.7 \times 10^{-6}$) | $\Gamma(5, 2.9 \times 10^{-7})$<br>$1.5 \times 10^{-6}$ ($4.7 \times 10^{-7}$, $2.9 \times 10^{-6}$) | $\Gamma(5, 5.2 \times 10^{-7})$<br>$2.6 \times 10^{-6}$ ($8.4 \times 10^{-7}$, $5.3 \times 10^{-6}$) | $\Gamma(5, 7.6 \times 10^{-7})$<br>$3.8 \times 10^{-6}$ ($1.2 \times 10^{-6}$, $7.8 \times 10^{-6}$) | $\Gamma(5, 4.9 \times 10^{-7})$<br>$2.5 \times 10^{-6}$ ($8.0 \times 10^{-7}$, $5.0 \times 10^{-6}$) | $\Gamma(5, 3.1 \times 10^{-7})$<br>$1.6 \times 10^{-6}$ ($5.1 \times 10^{-7}$, $3.2 \times 10^{-6}$) |
|  |  |  | Posterior: |  | 2.1e-06 (1.4e-06, 2.7e-06) | 1.7e-06 (1.3e-06, 2.2e-06) | 1.5e-06 (1.1e-06, 1.9e-06) | 2.7e-06 (2.1e-06, 3.1e-06) | 3.8e-06 (3.1e-06, 4.7e-06) | 1.8e-06 (1.3e-06, 2.2e-06) | 1.5e-06 (1.1e-06, 1.8e-06) |

| $m_{CHR}$ | Transmission rate among care home residents | - | Regional Prior | | $\Gamma(5, 4.3 \times 10^{-7})$ | $\Gamma(5, 3.7 \times 10^{-7})$ | $\Gamma(5, 2.9 \times 10^{-7})$ | $\Gamma(5, 5.2 \times 10^{-7})$ | $\Gamma(5, 7.6 \times 10^{-7})$ | $\Gamma(5, 4.9 \times 10^{-7})$ | $\Gamma(5, 3.1 \times 10^{-7})$ |
| --- | --- | --- | --- | --- | --- | --- | --- | --- | --- | --- | --- |
| | | | | | $2.2 \times 10^{-6}$ ($7.0 \times 10^{-7}$, $4.4 \times 10^{-6}$) | $1.8 \times 10^{-6}$ ($5.9 \times 10^{-7}$, $3.7 \times 10^{-6}$) | $1.5 \times 10^{-6}$ ($4.7 \times 10^{-7}$, $2.9 \times 10^{-6}$) | $2.6 \times 10^{-6}$ ($8.4 \times 10^{-7}$, $5.3 \times 10^{-6}$) | $3.8 \times 10^{-6}$ ($1.2 \times 10^{-6}$, $7.8 \times 10^{-6}$) | $2.5 \times 10^{-6}$ ($8.0 \times 10^{-7}$, $5.0 \times 10^{-6}$) | $1.6 \times 10^{-6}$ ($5.1 \times 10^{-7}$, $3.2 \times 10^{-6}$) |
| Posterior: |  |  |  |  | 2.2e-06 (1e-06, 3.4e-06) | 2.5e-06 (1.4e-06, 3.6e-06) | 1.6e-06 (7e-07, 2.4e-06) | 3.4e-06 (2.1e-06, 4.3e-06) | 2.8e-06 (5e-07, 4.8e-06) | 4.2e-06 (3.8e-06, 4.6e-06) | 3.3e-06 (2.9e-06, 3.6e-06) |
| $p_H^{max}$ | Probability of hospitalisation if symptomatic | $\psi_H^i$ | B(15.8, 5.28) | 0.75 (0.55, 0.91) | 0.87 (0.8, 0.92) | 0.9 (0.85, 0.94) | 0.89 (0.83, 0.95) | 0.78 (0.73, 0.84) | 0.85 (0.79, 0.9) | 0.86 (0.81, 0.93) | 0.73 (0.68, 0.79) |
| $p_{G_D}^{max}$ | Probability of death in care home if requiring hospitalisation | $\psi_{G_D}^i$ | B(15.8, 5.28) | 0.75 (0.55, 0.91) | 0.66 (0.37, 0.85) | 0.77 (0.64, 0.88) | 0.53 (0.41, 0.69) | 0.58 (0.52, 0.63) | 0.66 (0.5, 0.91) | 0.64 (0.6, 0.69) | 0.36 (0.32, 0.43) |
| $p_{ICU}^{max}$ | Probability of triage to ICU for new hospital admissions | $\psi_{ICU}^i$ | B(13.9, 43.9) | 0.24 (0.14, 0.36) | 0.15 (0.11, 0.18) | 0.15 (0.11, 0.18) | 0.17 (0.13, 0.21) | 0.25 (0.21, 0.31) | 0.31 (0.26, 0.37) | 0.12 (0.11, 0.13) | 0.23 (0.2, 0.25) |
| $p_{H_D}^{max}$ | Initial probability of death for general inpatients | $\psi_{H_D}^i$ | B(42.1, 50.1) | 0.46 (0.36, 0.56) | 0.42 (0.35, 0.5) | 0.46 (0.39, 0.53) | 0.43 (0.38, 0.47) | 0.47 (0.44, 0.51) | 0.37 (0.32, 0.46) | 0.5 (0.47, 0.53) | 0.41 (0.35, 0.46) |
| $p_{ICU_D}^{max}$ | Initial probability of death for ICU inpatients | $\psi_{ICU_D}^i$ | B(60.2, 29.3) | 0.67 (0.57, 0.77) | 0.66 (0.6, 0.72) | 0.71 (0.66, 0.77) | 0.69 (0.58, 0.77) | 0.69 (0.62, 0.75) | 0.61 (0.51, 0.69) | 0.71 (0.64, 0.77) | 0.63 (0.54, 0.75) |
| $p_{W_D}^{max}$ | Initial probability of death for stepdown inpatients | $\psi_{W_D}^i$ | B(28.7, 52.1) | 0.35 (0.25, 0.46) | 0.35 (0.25, 0.46) | 0.35 (0.3, 0.4) | 0.36 (0.3, 0.5) | 0.37 (0.3, 0.43) | 0.34 (0.24, 0.43) | 0.51 (0.44, 0.59) | 0.37 (0.29, 0.45) |
| $\mu_{ICU}$ | ICU admission multiplier after improvement in care | - | $U[0,1]$ | 0.5 (0.03, 0.98) | 0.79 (0.59, 0.93) | 0.76 (0.62, 0.93) | 0.72 (0.56, 0.94) | 0.51 (0.37, 0.64) | 0.62 (0.51, 0.75) | 0.83 (0.73, 0.99) | 0.44 (0.3, 0.54) |
| $\mu_D$ | Hospital mortality multiplier after improvement in care | - | $U[0,1]$ | 0.5 (0.03, 0.98) | 0.58 (0.45, 0.69) | 0.47 (0.41, 0.53) | 0.49 (0.42, 0.61) | 0.47 (0.42, 0.56) | 0.32 (0.27, 0.38) | 0.35 (0.28, 0.43) | 0.53 (0.44, 0.63) |
| $p_{NC}$ | Prevalence of non-COVID symptomatic leading to test | - | $U[0,1]$ | 0.5 (0.03, 0.98) | 0.0031 (0.0029, 0.0033) | 0.0022 (0.0021, 0.0024) | 0.0025 (0.0023, 0.0027) | 0.0028 (0.0026, 0.0031) | 0.0028 (0.0027, 0.003) | 0.0019 (0.0018, 0.002) | 0.003 (0.0028, 0.0031) |
| $\rho_{P2_{test}}$ | Overdispersion of PCR positivity | - | $U[0,1]$ | 0.5 (0.03, 0.98) | 0.0052 (0.0042, 0.0062) | 0.0076 (0.0064, 0.0086) | 0.0072 (0.0058, 0.0088) | 0.0033 (0.0029, 0.0037) | 0.0026 (0.0021, 0.0031) | 0.0091 (0.0079, 0.0103) | 0.0032 (0.0027, 0.0037) |

Age-specific scaling factors for each parameter are set out in Table S 8.\*\* Region codes: NW = North West, NEY = North East and Yorkshire, MID = Midlands, EE = East of England, LON = London, SW = South West, SE = South East. N.B. when the prior is region specific the prior is shown in the same columns as the posterior distributions

**Table S 7:** Fixed model parameters (age / care home scaling factors are shown separately in Table S 8).

| Parameter | Description | Value | Source |
| --- | --- | --- | --- |
| $p_c$ | Probability of developing symptoms after becoming infectious | 0.6 | Lavezzo et al. (44) |
| $p^*$ | Probability of arriving at hospital with a confirmed diagnosis | 0.25 | NHS (45) |
| $1/\gamma_U$ | Mean time to confirmation of diagnosis within hospital (days) | 2 days | CHES (5) |
| $p_{sero_{pos}}$ | Probability of seroconversion | 0.85 | Brazeau et al. (42) |
| $1/\gamma_{sero_{pos}}$ | Mean time to seroconversion from infectiousness (days) | 13 | Benny et al. (17) |
| $p_{sero_{spec}}$ | Specificity of serology test | 0.99 | Brazeau et al. (42) |
| $p_{sero_{sens}}$ | Sensitivity of serology test | 1 | Assumption as non-distinguishable from time varying seroconversion |

**Table S 8:** Age / care-home scaling factors

| Age / care home -stratified scaling to probability of: |  |  |  |  |  |  |
| --- | --- | --- | --- | --- | --- | --- |
| | Hospitalisation<br>if symptomatic ($p_H$) | Triage to ICU<br>($p_{ICU}$) | Death for general<br>inpatients ($p_{HD}$) | Death in ICU<br>($p_{ICUD}$) | Death in stepdown<br>($p_{WD}$) | Death in the<br>community ($p_{GD}$) |
| Population group | $\psi_H^i$ | $\psi_{ICU}^i$ | $\psi_{HD}^i$ | $\psi_{ICUD}^i$ | $\psi_{WD}^i$ | $\psi_{GD}^i$ |
| *Age* |  |  |  |  |  |  |
| [0, 5) | 0.039 | 0.243 | 0.039 | 0.282 | 0.091 | 0 |
| [5, 10) | 0.001 | 0.289 | 0.037 | 0.286 | 0.083 | 0 |
| [10, 15) | 0.006 | 0.338 | 0.035 | 0.291 | 0.077 | 0 |
| [15, 20) | 0.009 | 0.389 | 0.035 | 0.299 | 0.074 | 0 |
| [20, 25) | 0.026 | 0.443 | 0.036 | 0.310 | 0.074 | 0 |
| [25, 30) | 0.040 | 0.503 | 0.039 | 0.328 | 0.076 | 0 |
| [30, 35) | 0.042 | 0.570 | 0.045 | 0.353 | 0.080 | 0 |
| [35, 40) | 0.045 | 0.653 | 0.055 | 0.390 | 0.086 | 0 |
| [40, 45) | 0.050 | 0.756 | 0.074 | 0.446 | 0.093 | 0 |
| [45, 50) | 0.074 | 0.866 | 0.107 | 0.520 | 0.102 | 0 |
| [50, 55) | 0.138 | 0.954 | 0.157 | 0.604 | 0.117 | 0 |
| [55, 60) | 0.198 | **1.000** | 0.238 | 0.705 | 0.148 | 0 |
| [60, 65) | 0.247 | 0.972 | 0.353 | 0.806 | 0.211 | 0 |
| [65, 70) | 0.414 | 0.854 | 0.502 | 0.899 | 0.332 | 0 |
| [70, 75) | 0.638 | 0.645 | 0.675 | 0.969 | 0.526 | 0 |
| [75, 80) | **1.000** | 0.402 | 0.832 | **1.000** | 0.753 | 0 |
| 80+ | 0.873 | 0.107 | **1.000** | 0.918 | **1.000** | 0 |
| *Care home* |  |  |  |  |  |  |
| CHW | 0.104 | 0.784 | 0.134 | 0.519 | 0.114 | 0 |
| CHR | 0.873 | 0.107 | **1.000** | 0.918 | **1.000** | **1** |

## 2 Supplementary Results

### 2.1 Model fitting

**Figure S 6:**
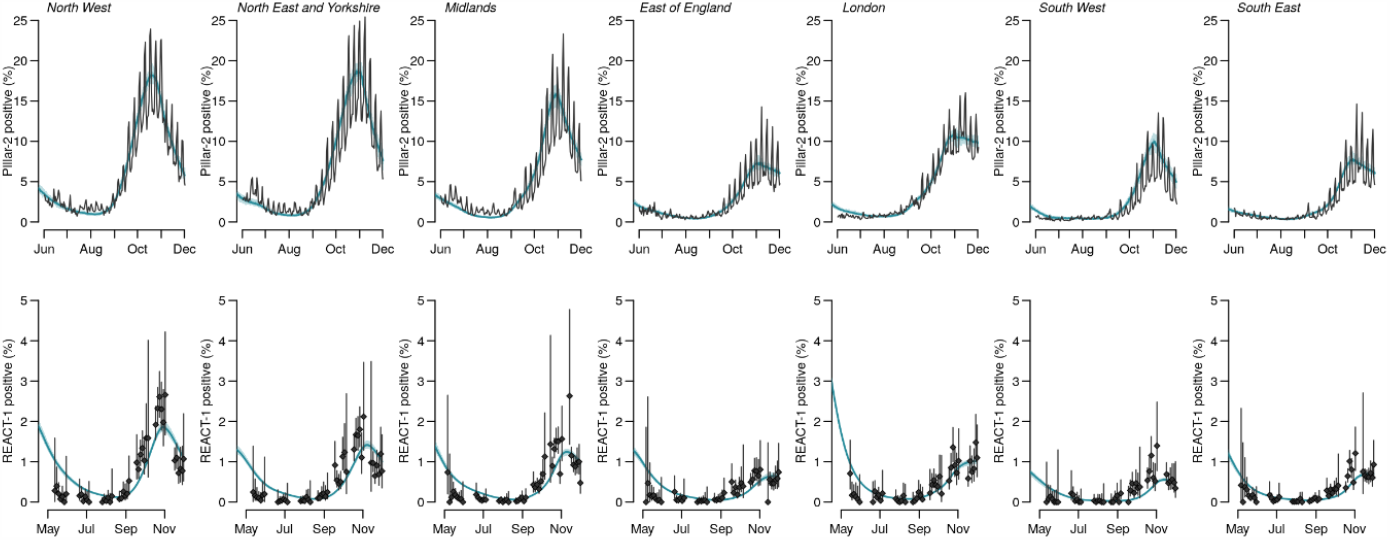
Model fits by region to PCR positivity for individuals aged >25 years (top row) and PCR positivity from the REACT-1 study (bottom row). The points show the data and bars the 95% CI. The solid line the median model fit and the shaded area the 95% CrI.

**Figure S 7:**
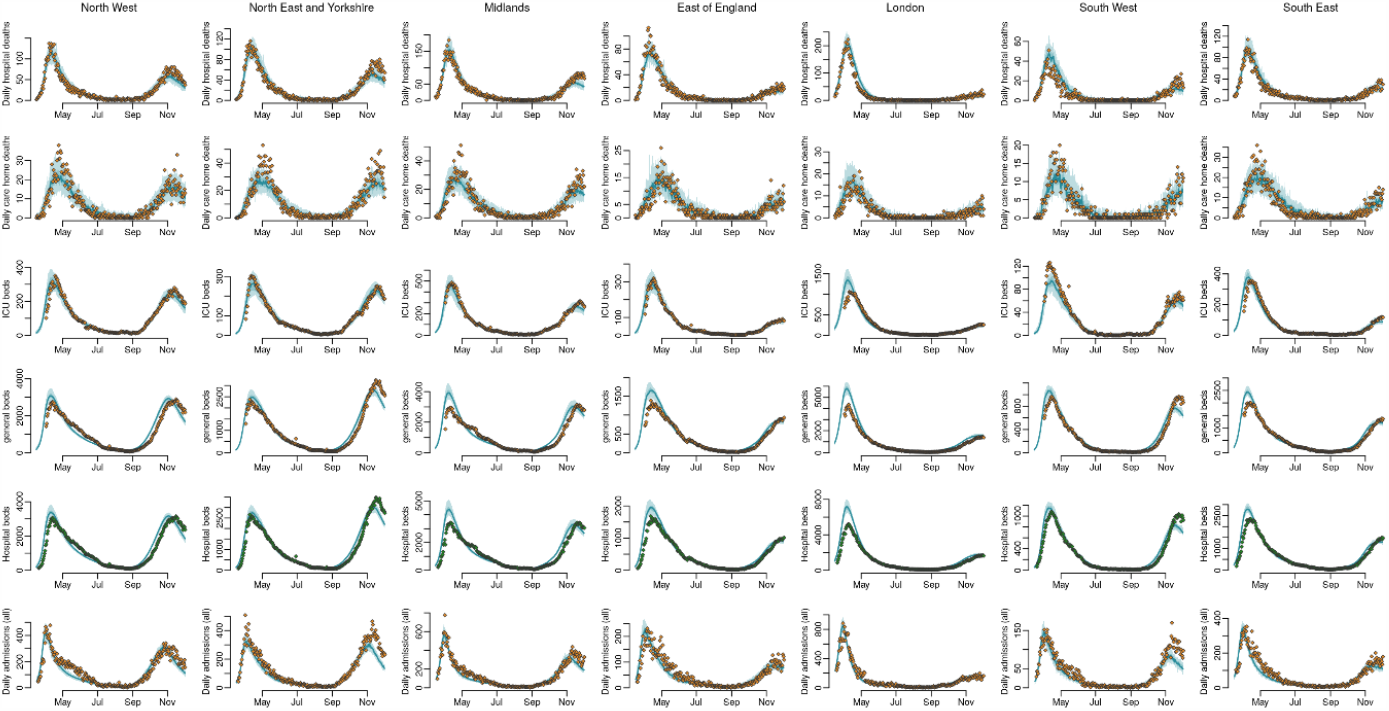
Model fits to daily hospital deaths (top row), daily care home deaths (second row), ICU bed occupancy (third row), general bed occupancy (fourth row), all hospital beds (fifth row), and all daily admissions (bottom row) by region (columns). The points show the data, the solid line the median model fit and the shaded area the 95% CrI.

### 2.2 Severity estimates

**Figure S 8:**
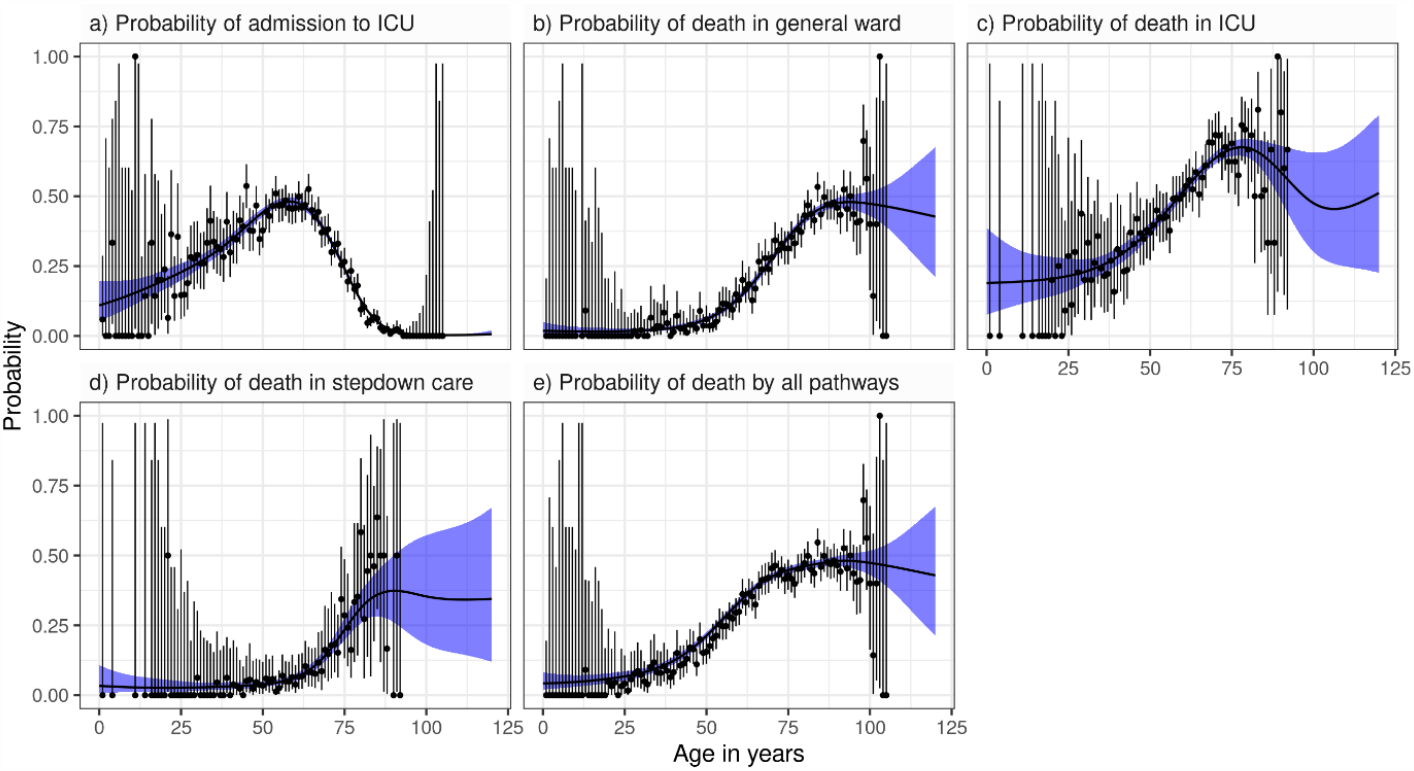
Fits to CHESS data broken down into one-year age bands. Blue ribbons show the 95% CrI of the fitted spline, black circles and vertical segments give the raw mean and 95% CI from the data (exact binomial).

**Table S 9:** Age-stratified estimates of disease severity (*to 2sf, ^to 3dp)

| Age stratified estimate of: |  |  |
| --- | --- | --- |
| Age group | Proportion of infections who were hospitalised* | Proportion of infections who died^ |
| [0, 5) | 3.0% (2.8%, 3.2%) | 0.031% (0.027%, 0.034%) |
| [5, 10) | 0.26% (0.24%, 0.28%) | 0.003% (0.002%, 0.003%) |
| [10, 15) | 0.084% (0.078%, 0.089%) | 0.001% (0.001%, 0.001%) |
| [15, 20) | 0.042% (0.039%, 0.045%) | 0.000% (0.000%, 0.001%) |
| [20, 25) | 0.080% (0.075%, 0.085%) | 0.001% (0.001%, 0.001%) |
| [25, 30) | 0.26% (0.24%, 0.28%) | 0.004% (0.003%, 0.004%) |
| [30, 35) | 0.40% (0.37%, 0.42%) | 0.006% (0.006%, 0.007%) |
| [35, 40) | 0.63% (0.58%, 0.67%) | 0.013% (0.011%, 0.014%) |
| [40, 45) | 1.2% (1.1%, 1.2%) | 0.031% (0.026%, 0.035%) |
| [45, 50) | 1.9% (1.8%, 2.1%) | 0.070% (0.061%, 0.080%) |
| [50, 55) | 2.3% (2.2%, 2.5%) | 0.116% (0.101%, 0.133%) |
| [55, 60) | 4.0% (3.8%, 4.3%) | 0.276% (0.242%, 0.315%) |
| [60, 65) | 9.6% (8.9%, 10%) | 0.867% (0.762%, 0.971%) |
| [65, 70) | 10% (9.6%, 11%) | 1.215% (1.070%, 1.352%) |
| [70, 75) | 24% (22%, 26 %) | 3.512% (3.083%, 3.900%) |
| [75, 80) | 50% (46%, 53%) | 8.430% (7.407%, 9.338%) |
| 80+ | 50% (47%, 54%) | 9.696% (8.501%, 10.640%) |
| Combined | 20% (13%, 27%) | 34.132% (28.020%, 41.359%) |

## Supplementary counterfactual analysis

**Figure S 9:**
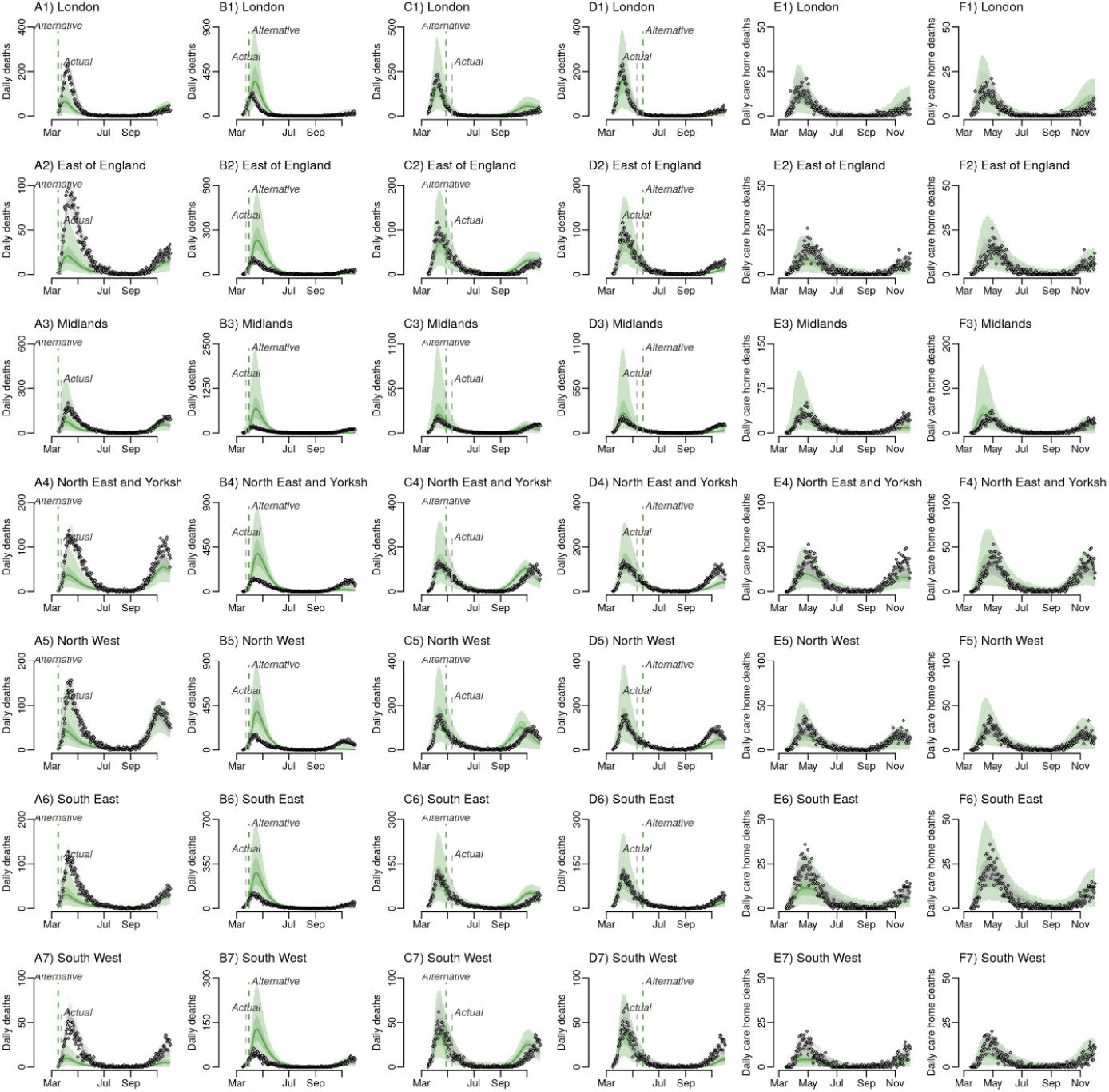
Counterfactual intervention scenarios in each England NHS Region: Panel **A1**-7 impact of locking down one-week earlier Panel **B**1-7 impact of locking down one week later; Panel **C**1-7 impact of relaxing lockdown restrictions two weeks earlier. Panel **D**1-7 impact of relaxing lockdown restrictions two weeks later; Panel **E**1-7 impact of 50% less contact between care home residents and the general population; Panel **F**1-7 impact of 50% more contact between care home residents and the general population.

## Notes

### Competing Interest Statement

The authors have declared no competing interest.

### Author Declarations

The use of pillar 2 PCR testing data was made possible thanks to PHE colleagues. The use of serological data was made possible by colleagues at PHE Porton Down, Colindale, and the NHS Blood Transfusion Service.

